# Improved screening of COVID-19 cases through a Bayesian network symptoms model and psychophysical olfactory test

**DOI:** 10.1101/2021.01.18.21249821

**Authors:** Susana Eyheramendy, Pedro A. Saa, Eduardo A. Undurraga, Carlos Valencia, Carolina López, Luis Méndez, Javier Pizarro-Berdichevsky, Andrés Finkelstein-Kulka, Sandra Solari, Nicolás Salas, Pedro Bahamondes, Martín Ugarte, Pablo Barceló, Marcelo Arenas, Eduardo Agosin

**Author notes:** S.E. and P.A.S. contributed equally to this work.

## Abstract

The infectiousness and presymptomatic transmission of SARS-CoV-2 hinder pandemic control efforts worldwide. Therefore, the frequency of testing, accessibility, and immediate results are critical for reopening societies until an effective vaccine becomes available for a substantial proportion of the population. The loss of sense of smell is among the earliest, most discriminant, and prevalent symptoms of COVID-19, with 75-98% prevalence when clinical olfactory tests are used. Frequent screening for olfactory dysfunction could substantially reduce viral spread. However, olfactory dysfunction is generally self-reported, which is problematic as partial olfactory impairment is broadly unrecognized. To address this limitation, we developed a rapid psychophysical olfactory test (KOR) deployed on a web platform for automated reporting and traceability based on a low-cost (about USD 0.50/test), six-odor olfactory identification kit. Based on test results, we defined an anosmia score –a classifier for olfactory impairment–, and a Bayesian Network (BN) model that incorporates other symptoms for detecting COVID-19. We trained and validated the BN model on two samples: suspected COVID-19 cases in five healthcare centers (*n* = 926; 32% COVID-19 prevalence) and healthy (asymptomatic) mining workers (*n* = 1, 365; 1.1% COVID-19 prevalence). All participants had COVID-19 assessment by RT-PCR assay. Using the BN model, we predicted COVID-19 status with 76% accuracy (AUC=0.79 [0.75 − 0.82]) in the healthcare sample and 84% accuracy (AUC=0.71 [0.63 − 0.79]) among miners. The KOR test and BN model enabled the detection of COVID-19 cases that otherwise appeared asymptomatic. Our results confirmed that olfactory dysfunction is the most discriminant symptom to predict COVID-19 status when based on olfactory function measurements. Overall, this work highlights the potential for low-cost, frequent, accessible, routine testing for COVID-19 surveillance to aid society’s reopening.

---

The COVID-19 pandemic has imposed an enormous toll, with more than 85 million cases and 1.9 million deaths globally as of January 2021 [1]. Despite recent effective COVID-19 vaccine approvals [2, 3, 4], epidemic containment will critically depend on non-pharmaceutical interventions (e.g., lockdowns, gathering restrictions) until a substantial proportion of the population is vaccinated [5, 6, 7]. These society-wide non-pharmaceutical strategies are socially and economically costly [8, 9]. As countries reopen and lift restrictions, there is a high risk of a resurgence of the epidemic [10, 11, 12]. More focused interventions are becoming essential to control viral transmission while reducing social and economic impact [13, 14, 15]. Controlling these transmission hotspots and effectively breaking the chain of viral transmission requires complementing non-pharmaceutical interventions with robust surveillance [16, 17].

Two characteristics of SARS-CoV-2, the virus that causes COVID-19, makes frequent screening, rapid diagnosis, and early isolation of infected individuals critical. First, the virus spreads efficiently, with an average number of secondary cases caused by an infected individual of about 2.5 [18, 19]. Second, a substantial proportion of onward transmission occurs before symptoms are apparent [20, 21, 22]. The viral load is the major spreading factor. It remains low during incubation time and reaches a peak slightly before symptoms onset [20]. This peak in infectiousness is followed by a rapid decline within about a week [20]. These two characteristics hinder epidemic control efforts because detection and isolation of infectious individuals are challenging. So far, COVID-19 surveillance has been mainly based on reverse transcription-polymerase chain reaction (RT-PCR) assays, considered the gold standard for diagnosis [23, 24]. Yet, RT-PCR assays are expensive and turnaround time, about 24 − 48 hours or longer, make them impractical as a surveillance tool to curb community transmission. Modeling studies have shown that effective COVID-19 surveillance should prioritize the frequency of testing, accessibility, and immediate results [16, 25, 26]. Infectious individuals might then be able to isolate and stop onward transmission promptly.This could be achieved, for example, using antigen tests which can have a turnaround time as low as 15 minutes and may cost about USD5-50 per test, or using mass screening for specific high-prevalence symptoms of SARS-CoV-2 infection, such as olfactory dysfunction [27].

Another critical aspect of a robust surveillance is having a clear characterization of the clinical presentation of COVID-19. Clinical signs and symptoms related to COVID-19 are mostly nonspecific and include cough, fever, shortness of breath, dyspnea, myalgia, and fatigue [28, 29, 30, 31]. Initially overlooked, the sudden loss of smell has emerged as one of the earliest and most prevalent symptoms of COVID-19 [32, 33, 34, 35, 36, 37, 38, 39]. The mechanisms that explain the loss of smell probably relate to an inflammatory response of support and vascular cells that could affect odor conduction by obstructing the olfactory clefts [40] or modifying olfactory sensory or olfactory bulb neurons’ function [41]. The function of olfactory sensory or olfactory bulb neurons could also be altered by damage to support cells[42] or vascular damage [43]. Recent studies have identified molecular factors involved in the sudden loss of smell [44]. SARS-CoV-2 uses the angiotensin-converting enzyme 2 (ACE2) and protease TMPRSS2 receptors to invade host cells [41]. Both proteins are expressed in various cell types and are particularly abundant in the nose, throat, upper bronchial airways, and alveolar epithelial type II cells. Protein expression in the nose has only been determined in supporting cells and stem cells in the olfactory epithelium, but not in olfactory neuronal receptors directly responsible for smell [40, 45]. These results suggest inflammation of cells in the olfactory epithelium leads to early loss and disturbance of the sense of smell in COVID-19 patients. Long-term symptoms could also be related to more extensive neural injury or virus persistence in the olfactory bulb [46, 47, 48]. However, olfactory dysfunction is seldom self-recognized and reported except by patients with the most severe smell disorders [49, 50, 51]. Therefore, self-reported partial (hyposmia) and total (anosmia) olfactory impairment associated with COVID-19 infection may be unreliable and shows substantial variation [38, 52, 53, 54, 55, 56]. In contrast, the prevalence of olfactory impairment in COVID-19 patients is high (75-98%) based on clinical psychophysical tests [57, 58, 59, 60, 61].

We have developed a model-based COVID-19 screening framework using a Bayes network symptom model and a low-cost (about USD 0.50/test) olfactory function test (KOR, Kit Olfativo Rápido) for frequent and immediate prediction of COVID-19 status. To support mass testing, we have deployed a secure web platform to store and track participants’ health state with automated reports. We present validation results of the Bayesian network model incorporating KOR test measurements on a sample of suspect COVID-19 patients (*n* = 926, 32% prevalence of SARS-CoV-2 infection) and asymptomatic healthy mining workers (*n* = 1365, 1.1% prevalence of infection). All participants had SARS-CoV-2 infection confirmation by RT-PCR assays. The KOR test and the results gathered in the platform can provide critical support for routine mass screening of COVID-19 symptoms for a safer and gradual reopening of society.

## Results

### KOR test and data

The KOR test is a standardized six-odor, forced multiple-choice identification test. Briefly, each participant is presented with six familiar odors, such as orange or vanilla, in a random sequence, one at a time. After the recognition of an odor, the individual is asked to select the term that best describes it from four options in a tablet or mobile phone (Figure 1A). Details of the KOR test design as well as the application protocol can be found in the corresponding *Methods* sections. After each identification, the selected choice is stored and used for model training using RT-PCR results as indicators of COVID-19 status. Figure 1B shows the proportion of individuals who recognized aromas, by RT-PCR status. 79−93% of participants with a negative RT-PCR recognized all aromas and 56 − 74% of participants with a positive RT-PCR recognized all aromas.

**Figure 1:**
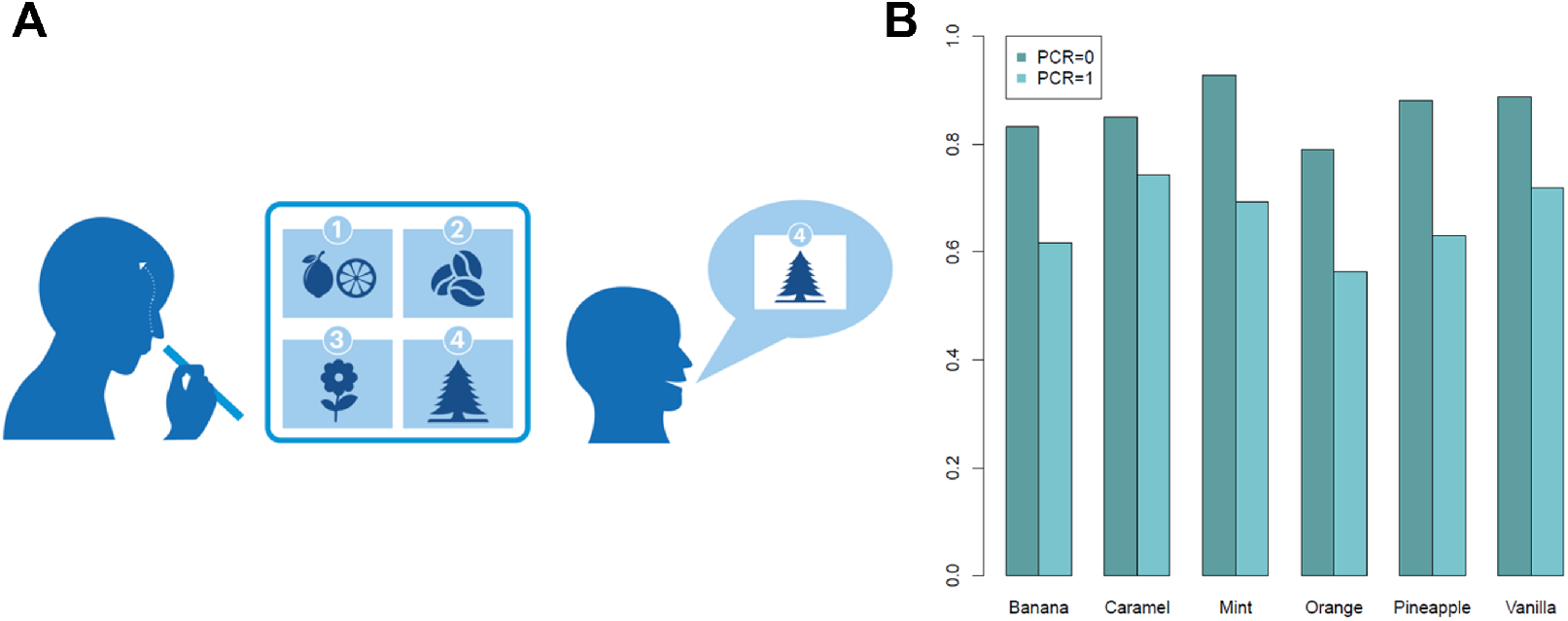
Implementation of the KOR test. (A) The KOR test consists of presenting individuals with six familiar odors, such as orange or vanilla, in a disposable piece of paper. Odors are presented in a random sequence. For each trial, we asked individuals to select in a tablet or mobile phone the term that best describes the odor from four options presented. If the individual does not recognize the odor, he/she selects “I do not recognize the odor”. The test implementation takes less than three minutes per participant. (B) Proportion of individuals from the UC-Christus Sample (*n* = 936) with a positive or negative result from the RT-PCR assay who perceived each aroma: Banana, Caramel, Mint, Orange, Pineapple and Vanilla.

We used two samples for the analysis. First, we gathered data from 926 patients with suspected COVID-19 infection or close contact with a laboratory-confirmed case in five medical centers in Santiago, Chile (UC-Christus). Our second sample consisted of 1, 365 healthy asymptomatic participants from a large mining operation in Chile, all of which underwent an exhaustive epidemiological and clinical screening before the KOR test. Participants in this sample were excluded from the study if they had any COVID-19 related symptom (cough, fever, shortness of breath, dyspnea, myalgia, and fatigue) or had been in contact with a lab-confirmed COVID-19 case in the past week. All participants in the “UC-Christus” and “miners” sample underwent an RT-PCR test following the KOR test. We considered all RT-PCR positive results as lab-confirmed COVID-19 [23, 24]. Descriptive statistics of the training sample, COVID-19 symptoms prevalence and RT-PCR status can be found in Supplementary Table S8.

### Anosmia score and classifier training

We developed an anosmia score based on the odors each participant was able to identify (see *Methods*). A higher score means a better sense of smell. Figure 2 compares anosmia score by RT-PCR status (Fig. 2A) and by self-reported anosmia (Fig. 2B), for all participants in the UC-Christus sample. Healthy individuals (RT-PCR negative) had a higher anosmia score than COVID-19 cases (Fig. 2A), and also showed higher anosmia scores than participants who reported normal sense of smell (Fig. 2B). The latter result suggests that some anosmic participants were not aware of their olfactory dysfunction, consistent with previous studies [56]. Participants with COVID-19 (RT-PCR positive) also showed a higher anosmia score than self-reported anosmic participants, suggesting there are normosmic participants among COVID-19 cases as reported elsewhere [39, 38].

**Figure 2:**
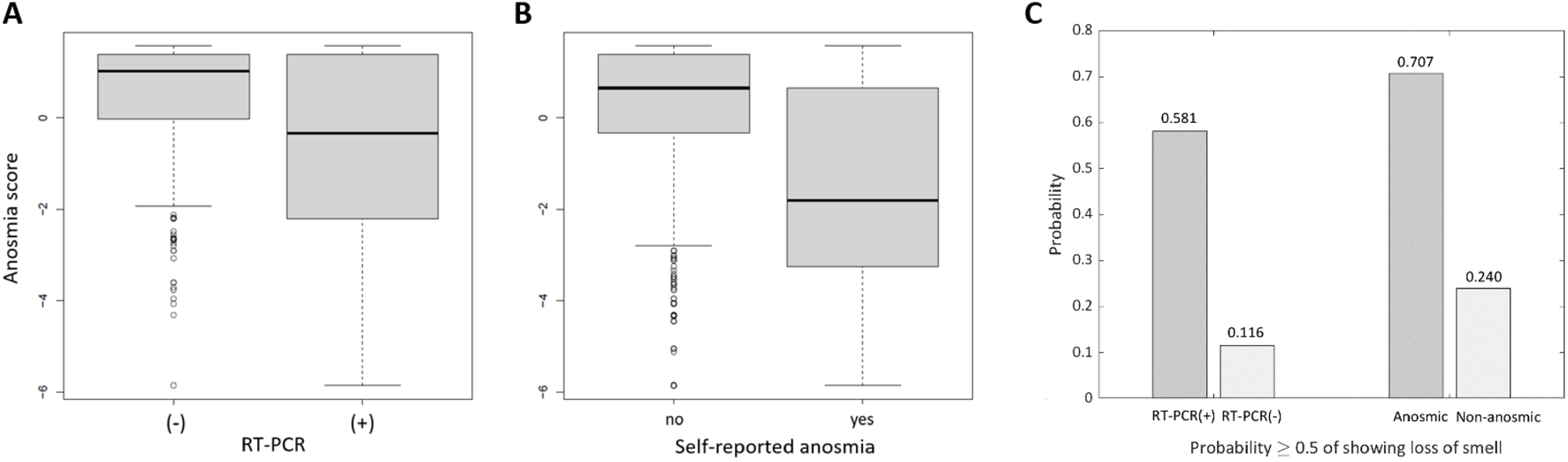
Anosmia score among participants. (A) Anosmia score distribution by RT-PCR status, and (B) by self-reported anosmia. Results include all UC-Christus participants (*n* = 926). (C) Results from applying a Gaussian mixture classifier to identify individuals with olfactory dysfunction.

Based on the anosmia scores we developed a Gaussian mixture classifier to identify individuals with olfactory impairment. This classifier is composed of two Gaussian distributions: one describes the anosmia scores of individuals with olfactory impairment, and the other represents the anosmia scores of individuals with a normal sense of smell.

A large proportion of COVID-19 cases develop some olfactory dysfunction [57, 58, 59, 62, 60, 61]. We estimated the distribution of the anosmia scores for the truly anosmic participants based on the scores obtained from self-reported anosmic participants (*mean* = −1.81; *std* = 2.1). Similarly, we estimated the distribution of the anosmia scores for the non-anosmic participants based on the score of the participants with negative RT-PCR ignoring outliers from the left tail (*mean* = 1.22; *std* = 0.97). We used these two distributions to build a Gaussian mixture classifier to identify individuals with olfactory dysfunction (refer to *Methods*). The results from the anosmia classifier are summarized in Figure 2C.

### Olfactory function measurement improves COVID-19 prediction by Bayesian network model in symptomatic population

We constructed a Bayesian network model to estimate whether a participant had COVID-19 (P) (Figure 3A). The model included self-reported cold (F) as a confounder, seven COVID-19 symptoms (cough, fever, muscular pain, breathing difficulty, self-reported anosmia, ageusia, and anosmia score), five indicator variables (sI1 recognized more than four odors; sI4 reported more than one symptom among cough, fever, breathing difficulty and muscular pain and sI1; sDays had 2-3 days with symptoms; sI7 had headache, diarrhea, or chest pain and recognized four or less odors; sI8 had ageusia, stomach pain, or fatigue, and sI1), and gender. This network structure was the most predictive out of several other models considering different subsets of symptoms and conditions (Supplementary Text S2, Table S1 and Figs. S1-S8). The contribution of each variable to the probability of having COVID-19 can be found in Supplementary Figs. S9-S10. For more details about the construction of the Bayesian network, the reader is referred to the corresponding subsection in *Methods*.

**Figure 3:**
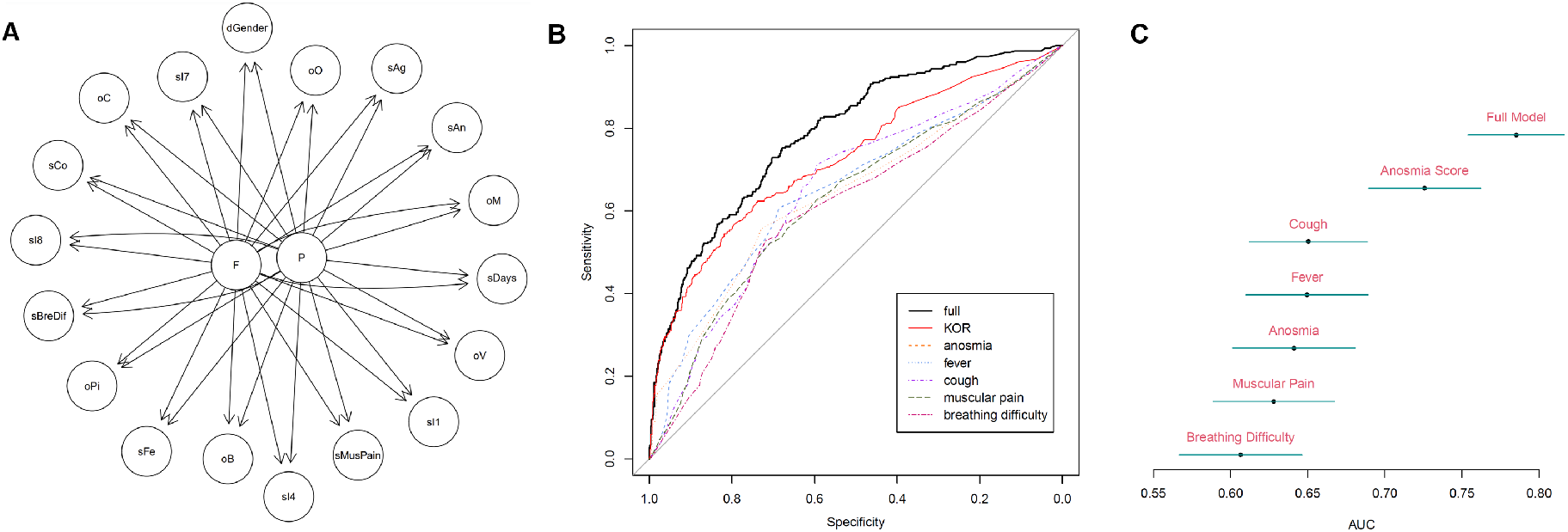
Training of the Bayesian network model for COVID-19 prediction. (A) Structure of the model. P represents the result of the RT-PCR assay, and F represents self-reported cold. The six different odors are represented with the variables: oB (Banana), oC (Caramel), oM (Mint), oO (Orange), oPi (Pineapple), oV (Vanilla). The six different symptoms are represented by: sCo (cough), sFe (fever), sMusPain (muscular pain), sBreDif (breathing difficulty), sAn (self-reported anosmia) and sAg (self-reported ageusia). Five indicator variables are represented by: sI1, sI4, sI7, sI8, sDays. dGender represents the gender of the individual. All variables are dichotomous. (B) Average Receiver Operating Characteristic (ROC) curve, as estimated through cross-validation (*K* = 10), for predicting a COVID-19 case using the full model (black line), only the KOR test (red line), and single self-reported symptoms. (C) Mean and 95% confidence intervals for the AUC of the ROC curves shown in (B). These analysis were performed using the UC-Christus sample (N=926)

We evaluated the performance of our model based on the UC-Christus dataset (*n* = 926, 32% positive RT-PCR). A 10-fold cross-validation was performed in order to have a robust estimator of the errors in the identification of positive and negative COVID-19 individuals. Figure 3B displays the average receiver operating characteristic (ROC) curve of the full model in black. To assess the contribution to the model of the objective measurements of olfactory impairment, we estimated the model with all variables except the ones that include information from the KOR test. Figure 3B shows the ROC curve of this model in red. The complete model displayed an area under the curve (AUC) of 0.785 with 95% confidence interval [0.754;0.816] and 76% accuracy (at a 0.5 threshold), whereas the partially complete model yielded an AUC of 0.733 with 95% confidence interval [0.696;0.768] and 72% accuracy (at a 0.5 threshold). The difference in the AUC is significant at an 87% level.

We further looked at the subset of individuals from the UC-Christus sample with no reported symptoms (*n* = 288) to assess model performance based solely on the anosmia score and measurements obtained from the KOR test. From this subset, 39 individuals had an RT-PCR positive, of which 9 had a predicted probability above 0.5 for being infected. All recognized three or less odors among a total of six, except for one that recognized four odors. The two odors not recognized were mint and orange. The remaining 30 individuals with a positive RT-PCR recognized at least four odors, and were not captured by the model at the infection probability threshold of 0.5. From the 249 individuals with a negative RT-PCR, 13 obtained a probability of being infected above 0.5, 11 of them recognized three or fewer odors and 2 recognized four, with mint among the non-recognized.

### Anosmia score predicts COVID-19 status with higher fidelity than self-report in asymptomatic cases

We further applied the KOR test to 1, 365 workers from a mining company that had not been previously infected and did not present any symptoms, such as fever, cough, and muscular pain. These workers were also tested with RT-PCR. Among all the workers, only 15 had a positive RT-PCR, of which the BN model identified 6. These six individuals identified less than four odors from the KOR test. The remaining nine individuals identified five out of the six odors (*n* = 4) or identified the six odors (*n* = 5). Overall, the model exhibited 96% accuracy, specificity 97% and sensitivity 40% at a 0.5 threshold (refer to Supplementary Table S7 for details.)

As the main predictor of COVID-19 status in asymptomatic individuals is their olfactory function, we compared the anosmia score of this sample and the probability of having olfactory dysfunction using the anosmia classifier trained with the UC-Christus dataset. The latter showed great agreement between the anosmia negative individuals and the miners cohort as observed by the overlapping between both densities (Figure 4A), confirming its suitability for describing the olfactory function of the general population. Finally, we compared the anosmia classifier results against the anosmia self-report of a sub-sample from the miners cohort (*n* = 825). There are 52 individuals likely of having olfactory dysfunction as determined by their anosmia score and the corresponding infection probability (*>* 0.5). This corresponds to 6.3% of the total sample. Of those, only 8 (approx. 1%) individuals self-reported an olfactory dysfunction. Roughly, 85% of individuals with olfactory impairment were not aware of their loss of the sense of smell (Figure 4B). This result confirms that olfactory impairment is typically underestimated or not perceived by healthy individuals [56], rendering the self-report of a “healthy olfactory function” unreliable.

**Figure 4:**
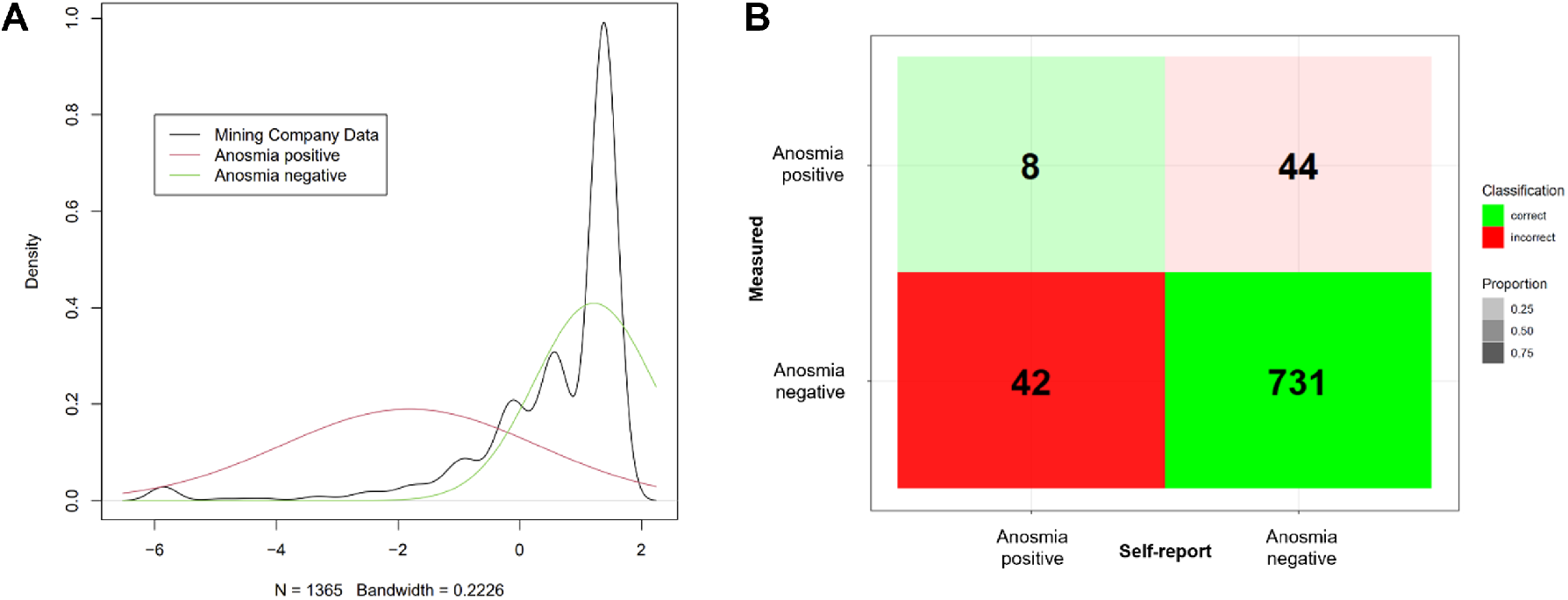
Anosmia prevalence in the healthy workers cohort. (A) Anosmia score density in the asymptomatic sample from the mining company (black line). The red and green lines correspond to the distribution for anosmic and non-anosmic individuals, respectively, as estimated in the UC-Christus data. (B) Comparison of self-reported anosmia and measured anosmia in the asymptomatic miners cohort. An individual was predicted to have olfactory dysfunction (anosmia) if the infection probability given its anosmia score was greater than 0.5.

## Discussion

We report a novel model for predicting the COVID-19 status of individuals based on self-reported symptoms (subjective measures) and the degree of olfactory dysfunction (anosmia score) as determined from psychophysical odor recognition trials from a rapid smelling test (Fig. 1). This anosmia score overcomes the known limitations of olfactory dysfunction self-report, i.e., usually underestimation of olfactory dysfunction severity (Fig. 2B), and furthermore, it substantially increases the predictive power of the model (Fig. 3B-C). We found that the full model incorporating this index yielded COVID-19 status predictions with high fidelity (AUC 0.785), which worsened when this index was left out (AUC 0.733). In the case of asymptomatic individuals (miners data), where the only measurement is obtained from the KOR test, the model showed a consistent performance. Among the 15 individuals that were RT-PCR positive, 6 of them recognized less than four odors, the remaining 9 recognized at least five out of the six different odors. We obtained a similar performance on the asymptomatic participants from the UC-Christus sample. The model recognized all individuals with RT-PCR positive who identified less than four odors and one individual who identified four odors. For the model to recognize an individual infected that identified four odors, mint has to be among the unrecognized odors.

As opposed to conventional olfactory function tests, KOR does not place the same weight on all the odors for the determination of the olfactory function state. A clear example is mint. Mint has two features that are especially attractive in a smell test. First, mint has a very distinctive smell and is rarely confused with other odors. Second, mint is a familiar smell to most people. In the KOR test, mint was correctly recognized by a majority of participants with a negative RT-PCR (93%). Hence, mint has a larger contribution to the model. If mint is not recognized there is a higher probability that the participant is infected (Fig. 1B). It is possible that some of the proposed odors may not be readily recognized across different sociocultural settings, despite being used and validated in international medical studies [63]. Cross-cultural validity would be, nevertheless, straightforward to address by piloting the test and by choosing odors which satisfy the two features mentioned above before implementation. Additional important aspects that distinguish KOR from other clinical olfactory tests are its simplicity, low cost, and speed. With these goals in mind, we devised an application protocol that was both easy to use (smell odorized paper strips) and of rapid application (15 seconds inter-stimulus time) (refer to *KOR test application protocol* subsection). It is likely that having such a simple and quick test may have affected tests results, and thus, the predictive capabilities of the model. For instance, typical inter-stimulus intervals are 40 seconds long, which allows to counteract the effect of external odorants (e.g., body fragrances) and also ease odor recognition. Careful design and verbal presentation of odor labels have also been shown to have an impact on odor recognition, particularly in normosmic individuals [64]. Optimization of the protocol thus constitutes an opportunity to further improve the test, and consequently, the predictive fidelity of the model.

We designed the KOR smell test and web platform as a low-cost public health tool for screening a large number of people, frequently, with a short sample-to-answer reporting time. The test’s purpose is to serve as an initial screening tool, leading to confirmation by a more accurate test such as a PCR or a reliable antigen test. Two aspects of SARS-CoV-2 infection make this epidemic difficult to control. First, the virus is efficiently transmitted between individuals, and transmissibility has increased in several places due to new circulating variants [65]. Second, a substantial portion of onward transmission occurs before symptoms are apparent, and the peak in infectiousness declines quickly, typically in about a week [66, 20, 67]. A substantial proportion of infected individuals do not have apparent symptoms [29, 68]. For example, the notorious Diamond Princess cruise ship had an asymptomatic COVID-19 infection prevalence of 30.8% in an adult population, and estimates suggest that about 4% of children are asymptomatic and 51% only have mild symptoms [69]. Several studies have stressed the importance of frequent, mass testing, with a short sample-to-answer reporting time to limit viral transmission in this epidemic [16, 17, 25, 26]. Several studies show that olfactory dysfunction is an early symptom [52, 54, 59, 70], but not necessarily the first to be apparent. Studies suggest that olfactory dysfunction is the first reported COVID-19 symptom in 12% [47], 27% [71], and 36% [72] of patients. However, because a large proportion of patients have olfactory dysfunction [47, 72, 73, 58, 33], several studies have highlighted the public health potential of screening for hyposmia or anosmia [74, 75, 27]. Modeling suggests that frequent screening for olfactory dysfunction could substantially reduce viral spread at a comparatively low cost [27]. Rational allocation of scarce diagnostic resources to test asymptomatic individuals is crucial, particularly in low- and middle-income countries. Despite the expectation that an olfactory test would be cost-effective [27], some settings with limited resources may not financially afford it. Until a substantial proportion of the population is vaccinated, frequent, accessible, routine screening for COVID-19 is critical to isolate infectious individuals and effectively break the chain of viral transmission. The KOR test, or any other psychophysical test to assess olfactory dysfunction would allow developing better sampling strategies that maximize the identification of high-probability COVID-19 cases with more sensitive tests (e.g., RT-PCR), among individuals with no apparent symptoms.

Our analyses are based on a sample of individuals from medical centers with probable COVID-19 and a sample of asymptomatic workers in a large mining operation. Neither sample is representative of the broader population. Nevertheless, the over-representation of positive cases in the UC-Christus data enabled us to estimate the relationship between symptoms and disease using a moderate sample size. The under-representation of positive cases in the miners data, challenged the model to capture infected individuals with just KOR test measurements. Furthermore, contexts like a mining operation are relevant as resemble a type of scenario where the test is more likely to be used as additional screening for individuals who do not have other apparent COVID-19 such as fever. The Bayesian network model has moderate accuracy and relatively low sensitivity. Diagnosis for symptomatic patients in a clinical setting usually requires high accuracy and sensitivity, as it defines a patient’s treatment. In contrast, given the transmission dynamics of SARS-CoV-2, rapid results and frequency of testing are much more critical for effective surveillance with the potential of controlling viral transmission [16, 17, 25, 26].

It is possible that false positives would be put under quarantine unnecessarily, at least until they can access a more sensitive test such as an RT-PCR. False positives may also occur because some Covid-19 patients have also reported persistent anosmia or hyposmia [46, **?**, 76, 77], and may test positive after the virus has been cleared. It is worth noting that patients may also show positive PCR results for weeks [78], with an average of 17 days [66] and a median of 22-33 days [79]. From an epidemiological standpoint, false positives are preferable to having community transmission, especially for a gradual reopening of society. Perhaps more worryingly, using a psychophysical smell test for mass screening would probably result in several false negatives and result in the onward transmission of SARS-CoV-2. However, in a counterfactual scenario without mass screening, those individuals would not be tested and still transmit the virus. The test may potentially lead to higher spreading if individuals without olfactory dysfunction but active infection felt falsely reassured. If individuals perceived the smell test as proof of no infection, they could weaken adherence to other prevention strategies such as wearing masks or social distancing. This compensatory increase in risky behavior when safety measures are in place, known as the “Peltzman Effect” [80], is partly determined by the perceived effectiveness of the measure and the motivation to take risky behaviors [81, 82]. Therefore, the potential use of this test requires adequate risk communication and is not intended to replace other non-pharmaceutical strategies for the control and prevention of viral transmission, such as social distancing, frequent hand-washing, and wearing face masks. Last, it is also possible that our test would fail to recognize some patients with olfactory dysfunction.

While they do not serve the same purpose, we also compared our test with standard RT-PCR results. Despite limited evidence, several studies have raised concerns about possible false-negative RT-PCR tests in patients with COVID-19 [83, 84, 85]. The evidence suggests that negative results, even with a relatively low probability of exposure, cannot rule out SARS-COV-2 infection [86]. Some of the cases we classified as false-positives due to negative RT-PCR results could have been true COVID-19 cases. This also underscores the importance of frequent testing for COVID-19 [16, 17, 27]. Similar to our smell test, antigen tests can have a sample-to-answer response time as low as 15 minutes. The performance of antigen tests varies substantially, and they are particularly susceptible to sampling quality [87]. For example, in the United Kingdom, the Innova test showed sensitivities for symptomatic patients of 79%, 73%, and 58% when used by trained laboratory scientists, trained healthcare staff, and pharmacy employees, respectively [88, 89]. While sensitivity is substantially lower in individuals without apparent symptoms [90, 12], rapid antigen tests can detect at least 66% of cases with high viral loads [12, 91]. At USD 5-50 per test, rapid antigen tests can substantially limit COVID-19 transmission through frequent mass testing. While the performance of our smell test is comparable to some antigen tests, our test is not intended for use as a diagnostic tool, but rather to serve as a low-cost (USD 0.50) psychophysical screening tool, leading to confirmation by a more accurate test.

Research among COVID-19 patients suggests that olfactory dysfunction, self-reported and based on a psychophysical test, was significantly more prevalent in mild compared to moderate-to-critical disease [46, 58]. The mechanisms that explain this observed difference are still unclear, but they could potentially be related to differences in patients’ immunological response by disease severity [46]. Olfactory loss may appear before other symptoms [92, 59] and seems to be more prevalent in mild compared to moderate-to-critical disease [47], which reinforces the potential of psychophysical smell test for early detection of COVID-19. An important extension of our study would be testing whether olfactory dysfunction allows distinguishing SARS-CoV-2 infections in patients with a negative RT-PCR test, which may be particularly problematic, for example, among healthcare and other essential workers (e.g., police). This could be addressed using serology, for example, testing individuals before the olfactory test and three weeks later.

Our model incorporated self-reported cold to capture the fact that several symptoms are shared between COVID-19 and cold. Nevertheless, several other diseases and conditions share symptoms with COVID-19, including smoking, allergies, and rhinitis, which we could have been considered in the model. However, capturing more complexity in the Bayesian network model requires an exponentially larger dataset, which rapidly becomes prohibitively expensive. We limited model complexity to make parameter estimation reliable. The model assumes that symptoms are independent given the status of cold and COVID-19 (infected/not infected). This independence assumption may not be satisfied, but we prioritized a simpler model that can capture the general tendency, given sample size constraints.

Until an effective vaccine for COVID-19 becomes available for a large proportion of the population, strategies to control the epidemic are mostly limited to non-pharmaceutical interventions. These interventions are socially and economically costly, and therefore, more focused interventions are increasingly important. Frequent screening, rapid diagnosis, and effective isolation of SARS-CoV-2 infections are critical. However, costs and time make currently available options impractical as a surveillance tool, particularly for low- and middle-income countries. We have presented a standardized, model-based, low-cost olfactory psychophysical test for the rapid screening of COVID-19. The test is painless and easy to implement and has a web-based secure platform for managing patient data. To this date (May 28, 2021), over 220,000 tests have been performed in the platform (refer to *KOR web platform*), which highlights the potential for systematic assessment of olfactory impairment in the general population. In a sample of participants with no apparent symptoms, the model captures individuals with loss of smell and a positive RT-PCR. Our results highlight the potential of using olfactory function assessments as a low-cost, frequent, accessible, painless, routine testing for COVID-19 surveillance for a safer reopening of society, including industry, universities, and other organizations.

## Methods

### COVID-19 RT-PCR test

Individuals were tested following the World Health Organization guidelines for real-time reverse-transcriptase PCR testing using validated diagnostics reported elsewhere [93].

### KOR test design

The KOR test is a rapid, six-odor, forced multiple-choice identification test. While previous rapid tests have shown to be reasonably effective for detecting total loss of olfactory function (i.e., anosmia) through the identification of only three odors [94], they are ineffective for the identification of partial olfactory function loss. As there is good evidence of COVID-19 cases suffering from partial olfactory impairment [58], we increased the number of odors in the test (6) to improve its sensitivity for detecting olfactory function loss. The proposed six odors were based on previous odor recognition reports [94] and clinical olfactory studies for infants in several countries including Chile [63]. To minimize recognition bias, candidate odors were evaluated and validated in a pilot trial. Only odors with a high recognition level of at least 85% (Supplementary Fig. S11) were selected. The sample of the pilot study consisted of 79% and 21% healthy men and women, respectively, with ages between 18 and 65 years of age in a sample of 2289 volunteers. Selected odors showed no significant differences in their recognition between men and women at 5% significance level (two-sided *t*-test, critical *p*-value = 0.21), displayed a high recognition level in all cases (*>* 85% one-sided *t*-test, minimum critical *p*-value = 0.41), and showed no significant differences in the average recognition level for individuals between 18 and 60 years at 5% significance level (ANOVA, critical *p*-value = 0.10). Details of the pilot study sample can be found in Supplementary Table S9.

### KOR test application protocol

The KOR test is a standardized, six-odor, forced multiple-choice identification test, which is managed through an online platform, hosted in www.testkor.com. The test is carried out as follows: a paper strip is impregnated with one drop (0.03 mL ± 4 uL) of an aromatic solution, as indicated by the platform, and presented to the participant to be smelled. The individual must identify the corresponding odour among 4 options, shown in the platform. Before evaluating the next odour, the subject is asked to neutralize eventual remaining fragrance in the nasal cavity by smelling his own body odor (wrist or forearm). All 6 aromas, i.e. banana, caramel, mint, orange, pineapple and vanilla, were evaluated in a random sequence, as determined by the platform, one at a time, with an inter stimulus interval of 15 seconds. The aromas were supplied by Alfa Group (https://alfagroup.cl/), a Chilean company specialized in providing food ingredients. The com-pany,s safety procedures are certified for HACCP and FSSC22000. All raw materials were FEMA / GRAS. Each aroma is contained in a 30 mL amber, glass dropper. Supplementary Table S10 indicates the composition and concentration of main odorants of the six solutions. The variability of the volume delivered by the glass dropper was measured in an analytical balance (*n* = 70), yielding a coefficient of variation lower than 15%, which is acceptable for odor recognition purposes. The reader is referred to the Supplementary Text S1 for more details on the application protocol.

### Model training dataset: COVID-19 suspects cohort from healthcare system

The first sample of participants was obtained from five medical centers that are part of the UC-Christus healthcare network in Santiago. The sample consisted of 926 individuals, 48% women and 52% men, with an average of 37 ± 13 years of age. These individuals had either symptoms compatible with SARS-CoV-2 infection, and/or were in close contact with confirmed cases. The individuals undergone both the olfactory KOR test and the SARS-CoV-2 real-time RT-PCR tests. COVID-19 prevalence was 32% in this sample. Individuals who reported base olfactory dysfunction as a consequence of previous trauma and/or acute/chronic health issues (e.g., chronic sinusitis, allergic rhinitis, among others) were not considered in this study. Before the application of the test, participants declared their COVID-19-related symptoms (fever, cough, muscular pain, breathing difficulty), demography (age, gender), prevailing diseases (allergy, cold, diabetes, hypertension, Parkinson, rhinitis and Alzheimer), and whether they were smokers. Other self-reported symptoms or conditions such as anosmia/hyposmia, ageusia, headache, diarrhea, fatigue, chest pain, and stomach pain, were also registered (refer to previous subsection).

### Model validation dataset: Workers cohort from mining company

For model validation under more realistic conditions, a sample of 1365 individuals was collected from a mining company in Antofagasta, Chile. Individuals that satisfy criteria i) and ii) (below) were simultaneously tested for their olfactory function status and SARS-CoV-2 real-time RT-PCR at a sanitary checkpoint. Inclusion criteria were: i) the person had not have a positive RT-PCR for COVID-19, and ii) at the moment of the test, the person did not have fever or any other apparent COVID-19 symptom. Only asymptomatic individuals were included in the sample. The prevalence of COVID-19 was 1.1% in this sample. Participants (825) also reported at the time of the screening if they recently suffered from olfactory impairment.

### Bayes network model

To predict an individual’s COVID-19 status, we built a model that considers an RT-PCR positive result as a COVID-19 case (*Y*_1_), and incorporates *cold* (*Y*_2_) as a possible confounder variable, seven symptoms (cough (*X*_7_), fever (*X*_8_), muscular pain (*X*_9_), breathing difficulty (*X*_10_), self-reported anosmia (*X*_14_), ageusia (*X*_15_) and the anosmia score), five indicator variables (*X*_11_, *X*_12_, *X*_13_, *X*_16_, *X*_17_) and gender (*X*_18_). To compute the anosmia score, we tested the identification of six odours: banana (*X*_1_), caramel (*X*_2_), mint (*X*_3_), orange (*X*_4_), pineapple (*X*_5_) and vanilla (*X*_6_). The five indicator variables were defined in the following way: *X*_11_ measures whether the individual recognized more than four odours or not (this score yielded the best COVID-19 prediction performance when the odors have equal weight, see Supplementary Fig. S13); *X*_12_ indicates whether the individual suffers from more than one symptom among cough, fever, muscular pain, breathing difficulty, headache, diarrhea and fatigue, and satisfies *X*_11_ = 1; *X*_13_ measures whether the individuals had 2-3 days with symptoms (this variable represents the days where the RT-PCR is most effective); *X*_16_ measures whether the individuals had *X*_11_ = 0 and one or more symptoms among headache, diarrhea and chest pain; and *X*_17_ measures whether the individual satisfies *X*_11_ = 1 and one or more symptoms among ageusia, stomach pain and fatigue. These indicator variables aim to identify group of symptoms that by occurring together increase the effect on the model. The structure of the model is depicted in Figure 3A. Note that the structure of the model assumes that all the X variables are conditionally independent given *Y*_1_ and *Y*_2_ and all X variables are colliders. The joint probability distribution of all variables is given by:

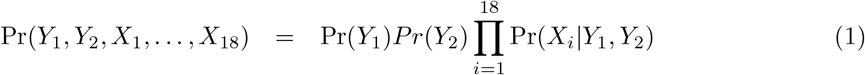

and we are interested in calculating the probability of positive RT-PCR given the other variables, i.e.

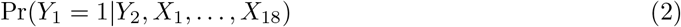

where all variables are dichotomous Bernoulli distributed. We assume a Binomial distribution for the joint probability of RT-PCR and self-reported cold.

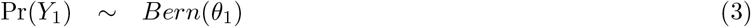

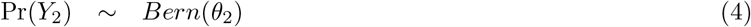

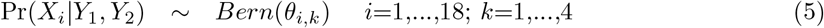

Here *k* denotes the four different outcomes that the joint variables (*Y*_1_, *Y*_2_) can take. Note that there are a total of 75 parameters that need to be estimated. *θ*_*k*_ represents the probability of belonging to group *k* for the variables *Y*_1_, *Y*_2_. *θ*_*i,k*_ represents the probability that an individual is positive for the condition/disease/variable *i* given that she/he is in group *k* for the variables *Y*_1_, *Y*_2_. We trained this model using a dataset of 926 individuals. In order to gain some degrees of freedom, and because we observed a clear tendency between the symptoms and the outcome of the variables *Y*_1_ and *Y*_2_ as shown in Supplementary Fig. S10, we set the following linear constrain on the parameters,

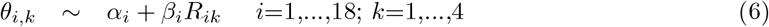

reducing the number of parameters to be estimated to 36. We either plug-in equations (3) and (4), or (3) and (5) into (1) to obtain via maximum likelihood the estimators of the parameters. The variables *R*_*ik*_ are known and set to fit the tendency of the parameters as shown in Supplementary Fig. S12.

Finally, for a new individual with variables *X*_1_ = *x*_1_, …, *X*_18_ = *x*_18_ we compute:

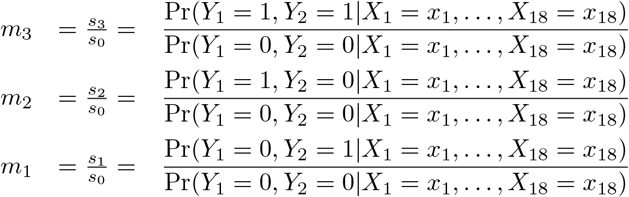

From these three equations we can obtain the probabilities *s*_0_, *s*_1_, *s*_2_, *s*_3_. If *Y*_2_ = 1 then

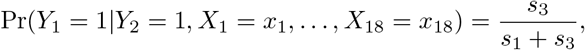

when *Y*_2_ = 0 then

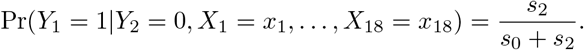

### Anosmia score and classifier

For an individual with outcomes from the KOR test given by (*x*_1_, *x*_2_, *x*_3_, *x*_4_, *x*_5_, *x*_6_) where *x*_*i*_ can take the value 1 if the individual identify odour *i* or 0 if the individual did not identify the odour *i*, its anosmia score is calculated in the following way.

- For individuals with the cold:

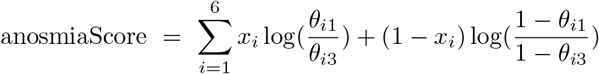
- For individuals without the cold:

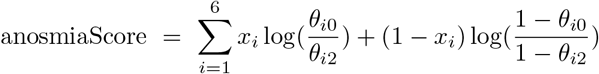

The distribution of the anosmia score, say *y*, is assumed to be a Gaussian mixture of two distribution:

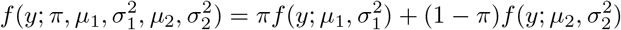

where *f* (*y*; *µ, σ*^2^) denotes a Gaussian distribution with mean *µ* and variance *σ*^2^, the parameters *µ*_1_, 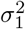 represent the parameters of the distribution for the anosmia score for individuals suffering from olfactory dysfunction, and the parameters *µ*_2_, 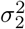 represent the parameters of the distribution for the anosmia score for individuals that do not suffer from olfactory dysfunction. The estimators for these parameters were obtained from the UC-Christus data: 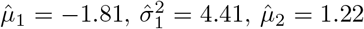 and 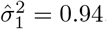

### KOR web platform

The KOR test web platform was designed to enable a rapid administration of the test. We developed a data model to store test information efficiently, paying attention to how often data about organizations, members, and screening tests are updated. The platform’s data access layer (back-end) was developed in Django (open-source framework), while its user interface (front-end) was developed in the JavaScript library ReactJS (open-source). High scalability and security concerns are handled by the deployment of the platform in Amazon Elastic Compute Cloud. General statistics on platform use, such as total tests, tests per day, and average time per test, can be found at http://metabase.imfd.cl/public/dashboard/2801acac-8414-43be-871d-dad441026d3a

## Data Availability

The required data to reproduce the results of this study have been deposited in Mendeley Data and can be freely downloaded from the link provided.

http://dx.doi.org/10.17632/z4ktvcwfp6.2

## Data availability

The required data to reproduce the results of this study have been deposited in Mendeley Data and can be freely downloaded at http://dx.doi.org/10.17632/z4ktvcwfp6.2

## Acknowledgements

This work was funded by the Technological Adoption Fund SiEmpre from SOFOFA Hub (CORFO), and partially funded by ANID through the Millennium Science Initiative Program ICN17 002 to S.E., P.B. and M.A.; ANID Millennium Science Initiative Program NCN17 081 and ANID/FONDAP CIGIDEN 15110017 to EU; FONDECYT 1200146 to S.E; and FONDECYT de Iniciación 11190871 to P.A.S. We would also like to acknowledge the support of the Center for Aromas and Flavors staff for helping in the collection of the data and Camila Pavesi for assisting in the organization and preliminary analysis of the latter.

## Contributions

S.E., P.A.S., E.A.U., L.M., J.P., A.F., s.s., P.B., M.A., and E.A. designed the study; P.A.S., C.V., C.L., and E.A. designed the olfactory test; C.V., C.L., and S.S. collected the data; S.E. performed modelling; M.U., N.S., and P.B. developed the web platform; S.E., P.A.S., E.A.U., and M.A. analyzed data; and S.E., P.A.S., E.A.U., M.A., and E.A. wrote the paper.

