## Supplementary Information for "Improved screening of COVID-19 cases through a Bayesian network symptoms model and psychophysical olfactory test"

[a]Faculty of Engineering and Science, Universidad Adolfo Ibáñez, Santiago, Chile
[b]Millennium Institute for Foundational Research on Data (IMFD), Santiago, Chile
[c]Department of Chemical and Bioprocess Engineering, School of Engineering, Pontificia Universidad Católica de Chile, Santiago, Chile
[d]Institute for Mathematical and Computational Engineering, Pontificia Universidad Católica de Chile, Santiago, Chile
[e]School of Government, Pontificia Universidad Católica de Chile, Santiago, Chile
[f]Millennium Initiative for Collaborative Research in Bacterial Resistance (MICROB-R), Santiago, Chile
[g]Research Center for Integrated Disaster Risk Management (CIGIDEN), Santiago, Chile
[h]Canadian Institute for Advanced Research (CIFAR), Toronto, Canada
[i]Center for Aromas and Flavors, DICTUC SA., Santiago, Chile
[j]Endoscopy Unit, Hospital Padre Hurtado, Santiago, Chile
[k]Department of Gastroenterology, Clinica Alemana de Santiago, Santiago, Chile
[l]Center for Innovation in Pelvic Floor, Hospital Sótero del Río, Santiago, Chile
[m]Department of Obstetrics and Gynecology, Faculty of Medicine, Pontificia Universidad Católica de Chile, Santiago, Chile
[n]Department of Otolaryngology, Clínica Alemana de Santiago, Santiago, Chile
[o]Faculty of Medicine, Clínica Alemana Universidad del Desarrollo, Santiago, Chile
[p]Department of Clinical Laboratory, Faculty of Medicine, Pontificia Universidad Católica de Chile, Santiago, Chile
[q]Department of Computer Science, School of Engineering, Pontificia Universidad Católica de Chile, Santiago, Chile
[*]S.E. and P.A.S. contributed equally to this work

This manuscript was compiled on May 31, 2021

### Supplementary Text

#### S1. KOR test protocol

To carry out an olfactory test, an evaluator must first enter the information of the individual to undergo the olfactory test. All this information is requested and stored in the KOR platform:

- Personal information: national identification number (or passport number), name, date of birth, nationality and gender.
- Information about previous SARS-CoV-2 test: the individual is asked whether he/she has undergone the SARS-CoV-2 RT-PCR test, and if the answer is yes, then he/she is asked the date of the test and whether the result was either negative or positive or still pending.
- Information on comorbidities: the individual is asked about his/her comorbidities. There is a predefined list that the evaluator can refer to, which includes: loss of smell, respiratory allergies, hypertension, rhinitis or sinusitis, Parkinson's disease, Alzheimer's disease, and diabetes. In addition, the platform provides an open question where the evaluator can enter any additional comorbidities.
- Information on smoking: the individual must indicate if he/she smokes.

This information is only requested once; the next time the individual undergoes the olfactory test, he/she only needs to provide the identification number in the platform for his/her information to be displayed.

As a second step, the evaluator must ask about the individual's symptoms and discomforts that may be associated with SARS-CoV-2. In particular, there is a check list that the evaluator has to use, which includes cough, cold or flu, fatigue, headache, and loss of appetite. If any symptom or discomfort is entered, then the number of days with this condition is asked. This data is requested every time an individual undergoes the olfactory test.

Next, the evaluator proceeds to ask six questions to measure partial or total loss of smell in the individual being tested. For each of these questions, the platform first tells the evaluator which odor (scent) to present to the individual, and then shows that person four pictures of odors asking him/her to identify the one he/she is smelling (see next section), and also giving a fifth alternative to indicate that he/she cannot identify it. It is important to notice here that the platform chooses at random the order in which the odors are presented, and also the distractors that will be used for each question.

Finally, according to the responses of the individual being tested, the platform produces a result in real time, which indicates if the individual has a reduction in his/her olfactory capacity, and therefore is a SARS-CoV-2 suspect.

##### Scents labels

Each scent of the KOR test is coded by a letter from "A" to "F", as follows: (A) Banana; (B) Caramel; (C) Mint; (D) Orange; (E) Pineapple; (F) Vanilla. Below, the available options for each multiple-choice odor recognition question are shown.

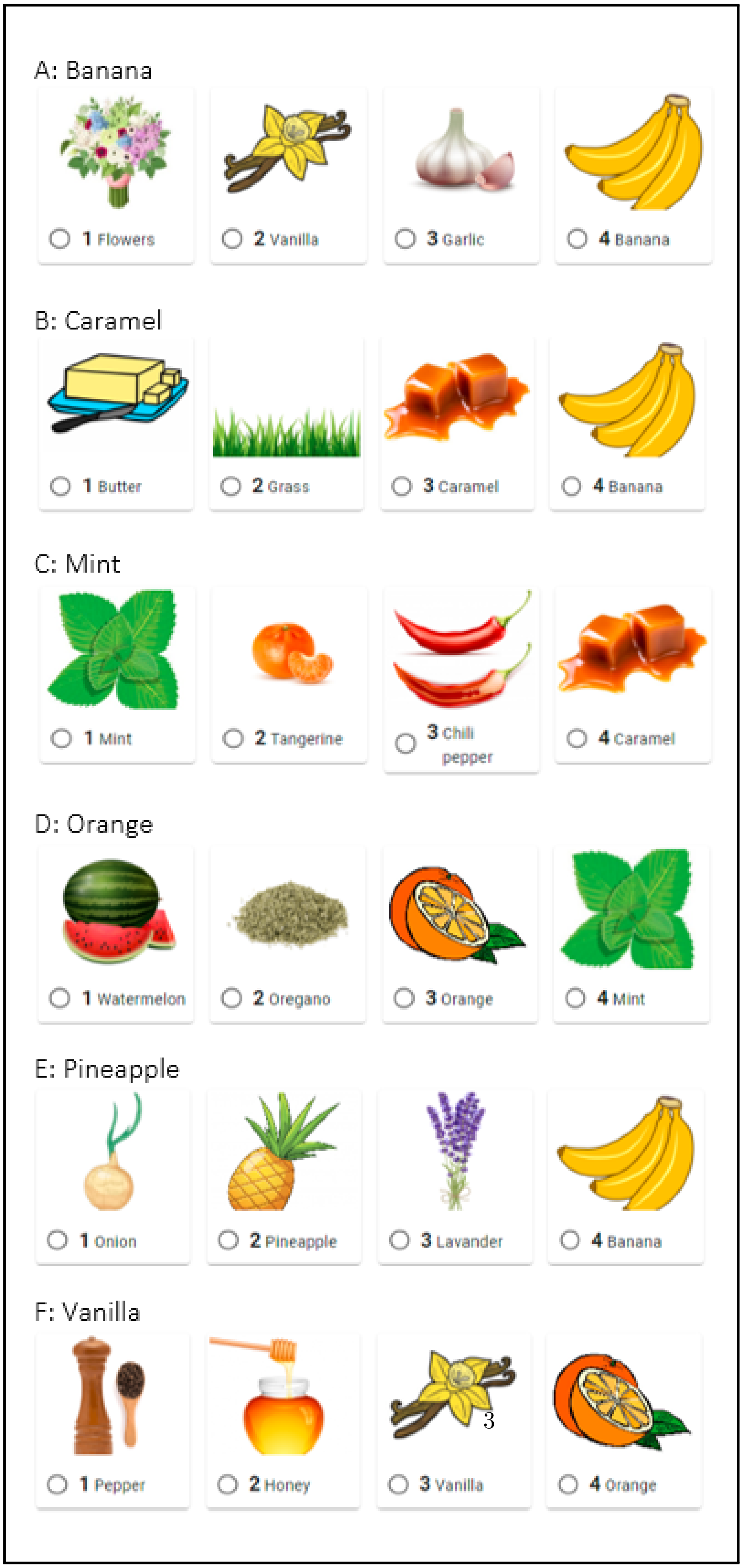
A: Banana
1 Flowers
2 Vanilla
3 Garlic
4 Banana
B: Caramel
1 Butter
2 Grass
3 Caramel
4 Banana
C: Mint
1 Mint
2 Tangerine
3 Chili pepper
4 Caramel
D: Orange
1 Watermelon
2 Oregano
3 Orange
4 Mint
E: Pineapple
1 Onion
2 Pineapple
3 Lavander
4 Banana
F: Vanilla
1 Pepper
2 Honey
3 Vanilla
4 Orange

#### S2. Model structure learning

In order to evaluate and identify the best structure for the Bayesian net, several network structures were assessed for their predictive performance. The variables considered in the models are described in what follows. P represents the result of the RT-PCR assay, F represents self-reported cold, R represents the presence or absence if rhinitis, A represents the presence or absence of allergies, and S whether the person smokes or not. These five variables P, F, A, S and R are considered either diseases or conditions that can lead to different symptoms which are the variables described next. The perception or not of six different odors are represented with the variables: oB (Banana), oC (Caramel), oM (Mint), oO (Orange), oPi (Pineapple), oV (Vanilla). Six different symptoms are represented by: sCo (cough), sFe (fever), sMusPain (muscular pain), sBreDif (breathing difficulty), sAn (self-reported anosmia), and sAg (self-reported ageusia). Additionally, five indicator variables are included and represented by (described in the *Methods* section of the main article): sI1, sI4, sI7, sI8, sDays. Finally, dGender represents the gender of the individual.

The evaluated structures are shown in Figures 1-6. These figures differ only on the diseases and/or conditions included in the model, whereas the remaining structures consider subsets of the symptoms Figs. 7-8. Tables 2, 3, 4, 5 and 6 show respectively the effect of the covariates for the dependent variables *allergies*, *rhinitis*, *cold*, *RT-PCR* and *smoking*, when a logistic regression model was fitted to the data. This model was employed as a starting point for identifying a smaller set of significant variables to be included in the final Bayesian Network model. Finally, Table 1 displays the performance of the evaluated network structures for the prediction of COVID-19 status in the training dataset.

#### Supplementary Tables

**Table 1:** Performance of different network structures for the prediction of COVID-19 status in the training dataset.

| Model structure | Estimated AUC* | 95% confidence interval AUC* | Reference |
|---|---|---|---|
| Final | 0.79 | [0.75 - 0.82] | Fig. 3 main text |
| 1 | 0.78 | [0.75 - 0.81] | Supplementary Fig. 1 |
| 2 | 0.77 | [0.73 - 0.80] | Supplementary Fig. 2 |
| 3 | 0.76 | [0.73 - 0.80] | Supplementary Fig. 3 |
| 4 | 0.77 | [0.74 - 0.80] | Supplementary Fig. 4 |
| 5 | 0.76 | [0.73 - 0.80] | Supplementary Fig. 5 |
| 6 | 0.76 | [0.73 - 0.80] | Supplementary Fig. 6 |
| 7 | 0.77 | [0.74 - 0.80] | Supplementary Fig. 7 |
| 8 | 0.76 | [0.73 - 0.80] | Supplementary Fig. 8 |

* These estimates were obtained by means of a 10-fold cross validation.

**Table 2:** Logistic regression results for rhinitis.

| Covariates | Estimate | Std. Error | $z$-value | Pr(>\|z\|) |
|---|---|---|---|---|
| (Intercept) | -1.17 | 0.74 | -1.59 | 0.11 |
| Banana | -0.24 | 0.27 | -0.89 | 0.37 |
| Caramel | 0.28 | 0.30 | 0.94 | 0.35 |
| Mint | 0.10 | 0.33 | 0.31 | 0.75 |
| Orange | -0.33 | 0.25 | -1.33 | 0.18 |
| Pineapple | -0.43 | 0.29 | -1.50 | 0.13 |
| Vanilla | -0.31 | 0.28 | -1.10 | 0.27 |
| Cough | 0.52 | 0.23 | 2.30 | 0.02 |
| Fever | -0.64 | 0.32 | -2.01 | 0.04 |
| sMusPain | 0.35 | 0.24 | 1.47 | 0.14 |
| sBreDif | -0.04 | 0.30 | -0.14 | 0.89 |
| Ageusia | -0.01 | 1.09 | -0.01 | 0.99 |
| Anosmia | 0.04 | 0.48 | 0.08 | 0.94 |
| sI1 | -0.17 | 0.41 | -0.42 | 0.68 |
| sI4 | -0.38 | 0.41 | -0.92 | 0.36 |
| sDays | 0.00 | 0.23 | 0.02 | 0.98 |
| sI7 | -0.21 | 0.32 | -0.64 | 0.52 |
| sI8 | -0.35 | 1.04 | -0.33 | 0.74 |
| dGender | -0.27 | 0.20 | -1.34 | 0.18 |

**Table 3:** Logistic regression results for smoking.

| Covariates | Estimate | Std. Error | $z$-value | Pr(>\|z\|) |
|---|---|---|---|---|
| (Intercept) | -1.54 | 0.60 | -2.55 | 0.01 |
| Banana | 0.00 | 0.21 | 0.01 | 0.99 |
| Caramel | -0.12 | 0.22 | -0.52 | 0.60 |
| Mint | 0.09 | 0.26 | 0.33 | 0.74 |
| Orange | 0.14 | 0.20 | 0.71 | 0.48 |
| Pineapple | 0.82 | 0.24 | 3.38 | 0.00 |
| Vanilla | -0.18 | 0.22 | -0.83 | 0.41 |
| Cough | -0.44 | 0.19 | -2.39 | 0.02 |
| Fever | -0.39 | 0.25 | -1.58 | 0.11 |
| sMusPain | -0.07 | 0.19 | -0.35 | 0.73 |
| sBreDif | -0.15 | 0.25 | -0.61 | 0.54 |
| Ageusia | -0.77 | 0.91 | -0.85 | 0.40 |
| Anosmia | 0.21 | 0.38 | 0.56 | 0.58 |
| sI1 | 0.38 | 0.32 | 1.21 | 0.22 |
| sI4 | 0.12 | 0.34 | 0.35 | 0.73 |
| sDays | 0.13 | 0.17 | 0.78 | 0.44 |
| sI7 | 0.05 | 0.24 | 0.22 | 0.83 |
| sI8 | 0.30 | 0.83 | 0.37 | 0.71 |
| dGender | 0.10 | 0.15 | 0.63 | 0.53 |

**Table 4:** Logistic regression results for allergies.

| Covariates | Estimate | Std. Error | $z$-value | Pr(>\|z\|) |
|---|---|---|---|---|
| (Intercept) | -2.53 | 0.85 | -2.98 | 0.00 |
| Banana | 0.19 | 0.30 | 0.64 | 0.53 |
| Caramel | 0.61 | 0.35 | 1.73 | 0.08 |
| Mint | 0.17 | 0.38 | 0.44 | 0.66 |
| Orange | -0.17 | 0.26 | -0.67 | 0.50 |
| Pineapple | -0.36 | 0.31 | -1.15 | 0.25 |
| Vanilla | 0.11 | 0.32 | 0.36 | 0.72 |
| Cough | 0.41 | 0.23 | 1.80 | 0.07 |
| Fever | -0.96 | 0.33 | -2.90 | 0.00 |
| sMusPain | 0.00 | 0.25 | 0.00 | 1.00 |
| sBreDif | 0.86 | 0.27 | 3.23 | 0.00 |
| Ageusia | 0.99 | 0.99 | 0.99 | 0.32 |
| Anosmia | 0.61 | 0.46 | 1.34 | 0.18 |
| sI1 | -0.53 | 0.47 | -1.12 | 0.26 |
| sI4 | 0.54 | 0.45 | 1.18 | 0.24 |
| sDays | -0.18 | 0.23 | -0.81 | 0.42 |
| sI7 | 0.08 | 0.30 | 0.26 | 0.79 |
| sI8 | -1.13 | 1.06 | -1.07 | 0.29 |
| dGender | 0.32 | 0.20 | 1.57 | 0.12 |

**Table 5:** Logistic regression results for cold.

| Covariates | Estimate | Std. Error | $z$-value | Pr(>\|z\|) |
|---|---|---|---|---|
| (Intercept) | -0.55 | 0.58 | -0.96 | 0.34 |
| Banana | -0.38 | 0.21 | -1.84 | 0.07 |
| Caramel | -0.06 | 0.22 | -0.26 | 0.79 |
| Mint | -0.43 | 0.25 | -1.70 | 0.09 |
| Orange | -0.53 | 0.20 | -2.69 | 0.01 |
| Pineapple | -0.23 | 0.22 | -1.02 | 0.31 |
| Vanilla | 0.02 | 0.23 | 0.07 | 0.94 |
| Cough | 0.84 | 0.17 | 4.84 | 0.00 |
| Fever | 0.64 | 0.21 | 3.00 | 0.00 |
| sMusPain | 0.62 | 0.18 | 3.42 | 0.00 |
| sBreDif | 0.07 | 0.22 | 0.32 | 0.75 |
| Ageusia | 1.34 | 0.92 | 1.46 | 0.15 |
| Anosmia | 0.34 | 0.36 | 0.94 | 0.35 |
| sI1 | -0.06 | 0.32 | -0.20 | 0.84 |
| sI4 | -1.12 | 0.32 | -3.54 | 0.00 |
| sDays | 0.86 | 0.17 | 4.93 | 0.00 |
| sI7 | -0.38 | 0.25 | -1.53 | 0.13 |
| sI8 | -1.38 | 0.88 | -1.57 | 0.12 |
| dGender | 0.11 | 0.16 | 0.70 | 0.49 |

**Table 6:** Logistic regression results for RT-PCR.

| Covariates | Estimate | Std. Error | $z$-value | Pr(>\|z\|) |
|---|---|---|---|---|
| (Intercept) | -0.14 | 0.66 | -0.21 | 0.83 |
| Banana | -0.49 | 0.22 | -2.22 | 0.03 |
| Caramel | 0.35 | 0.25 | 1.39 | 0.16 |
| Mint | -0.88 | 0.27 | -3.30 | 0.00 |
| Orange | -0.37 | 0.21 | -1.74 | 0.08 |
| Pineapple | -0.57 | 0.24 | -2.39 | 0.02 |
| Vanilla | -0.35 | 0.24 | -1.46 | 0.14 |
| Cough | 1.03 | 0.19 | 5.35 | 0.00 |
| Fever | 1.00 | 0.23 | 4.35 | 0.00 |
| sMusPain | 0.02 | 0.20 | 0.08 | 0.94 |
| sBreDif | -0.28 | 0.25 | -1.12 | 0.26 |
| Ageusia | -0.03 | 0.94 | -0.04 | 0.97 |
| Anosmia | 1.64 | 0.43 | 3.78 | 0.00 |
| sI1 | 0.53 | 0.34 | 1.56 | 0.12 |
| sI4 | -0.70 | 0.35 | -2.03 | 0.04 |
| sDays | 0.59 | 0.19 | 3.16 | 0.00 |
| sI7 | -0.62 | 0.29 | -2.17 | 0.03 |
| sI8 | 1.34 | 1.00 | 1.34 | 0.18 |
| dGender | 0.37 | 0.17 | 2.16 | 0.03 |

**Table 7:** Confusion matrix for the validation dataset.

| | Prob of COVID-19(+) | |
|---|---|---|
| RT-PCR | Negative ($p \leq 0.5$) | Positive ($p > 0.5$) |
| Negative | 1241 | 39 |
| Positive | 9 | 6 |

**Table 8:** Proportion of presence of symptoms and variables among PCR positive and PCR negative individuals.

| Variables | PCR negative | PCR positive |
|---|---|---|
| Cough | 0.27 | 0.51 |
| Fever | 0.10 | 0.31 |
| sMusPain | 0.32 | 0.48 |
| sBreDif | 0.12 | 0.18 |
| sI1 | 0.19 | 0.53 |
| sI4 | 0.06 | 0.23 |
| sDays | 0.44 | 0.68 |
| Anosmia | 0.02 | 0.16 |
| Ageusia | 0.00 | 0.08 |
| sI7 | 0.16 | 0.07 |
| sI8 | 0.00 | 0.09 |
| dGender | 0.51 | 0.53 |

**Table 9:** KOR test odor recognition statistics in the pilot study sample by different criteria

| Criteria | | | | Score | | | | Total |
|---|---|---|---|---|---|---|---|---|
| Genre | 0 | 1 | 2 | 3 | 4 | 5 | 6 | |
| Male | 4 | 15 | 23 | 58 | 169 | 520 | 1009 | 1798 |
| Female | 0 | 1 | 4 | 17 | 44 | 146 | 279 | 491 |
| Country of origin | | | | | | | | |
| Chile | 4 | 15 | 19 | 52 | 158 | 501 | 1083 | 1832 |
| Haití | 0 | 1 | 6 | 4 | 13 | 25 | 23 | 72 |
| Perú | 0 | 0 | 0 | 6 | 7 | 23 | 25 | 61 |
| Venezuela | 0 | 0 | 2 | 7 | 26 | 81 | 123 | 239 |
| Colombia | 0 | 0 | 0 | 4 | 5 | 14 | 17 | 40 |
| Other | 0 | 0 | 0 | 2 | 4 | 22 | 17 | 45 |
| Age range | | | | | | | | |
| 16-20 | 1 | 0 | 0 | 0 | 5 | 12 | 24 | 42 |
| 21-25 | 2 | 1 | 4 | 12 | 29 | 84 | 194 | 326 |
| 26-30 | 0 | 6 | 8 | 16 | 38 | 143 | 286 | 497 |
| 31-35 | 1 | 3 | 3 | 11 | 29 | 110 | 239 | 396 |
| 36-40 | 0 | 0 | 5 | 13 | 22 | 101 | 192 | 333 |
| 41-45 | 0 | 1 | 1 | 8 | 24 | 65 | 115 | 214 |
| 46-50 | 0 | 2 | 3 | 6 | 22 | 66 | 117 | 216 |
| 51-55 | 0 | 2 | 2 | 1 | 20 | 45 | 63 | 133 |
| 56-60 | 0 | 1 | 1 | 4 | 15 | 22 | 31 | 74 |
| 61-65 | 0 | 0 | 0 | 4 | 7 | 7 | 8 | 26 |
| 66-70 | 0 | 0 | 0 | 0 | 0 | 3 | 0 | 3 |

**Table 10:** Chemical composition of KOR test scents

| Scent | Provider code* | Solvent | Main odorant compounds | Concentration (g/L) |
|---|---|---|---|---|
| Banana | PTMEZ382401 | Propylene glycol | Isoamyl acetate | 11.49 |
| | | | Vanillin | 4.57 |
| | | | 2,3-Dihydroxypropyl acetate | 3.85 |
| | | | Phenethyl isovalerate | 3.50 |
| | | | Isoamyl butanoate | 3.00 |
| Caramel | PTMEZ382701 | Propylene glycol | Vanillin | 7.20 |
| | | | Ethyl maltol | 6.08 |
| | | | Nonanoic acid | 2.45 |
| | | | Furaneol | 1.57 |
| | | | Octanoic acid | 0.60 |
| Mint | PTMEZ381801 | Propylene glycol | Menthyl acetate | 14.40 |
| | | | p-Menthan-3-one | 3.80 |
| | | | D-Menthol | 2.10 |
| | | | D-Carvone | 2.00 |
| Orange | PTMEZ381901 | Tributyl acetylcitrate | D-Limonene | 2.60 |
| | | | Linalool | 1.70 |
| | | | 8-p-Menthadien-1,2-diol | 0.45 |
| | | | Decanal | 0.31 |
| Pineapple | PTMEZ382101 | Propylene glycol | Allyl hexanoate | 7.05 |
| | | | Isoamyl butanoate | 2.99 |
| | | | Ethyl hexanoate | 2.97 |
| | | | Ethyl octanoate | 1.40 |
| Vanilla | PTMEZ383177 | Propylene glycol | Ethyl isovanilline | 20.66 |
| | | | Benzyl alcohol | 4.67 |
| | | | Piperonal | 3.90 |
| | | | Vanillin | 0.63 |

* Alpha Group, Santiago, Chile.

#### Supplementary Figures

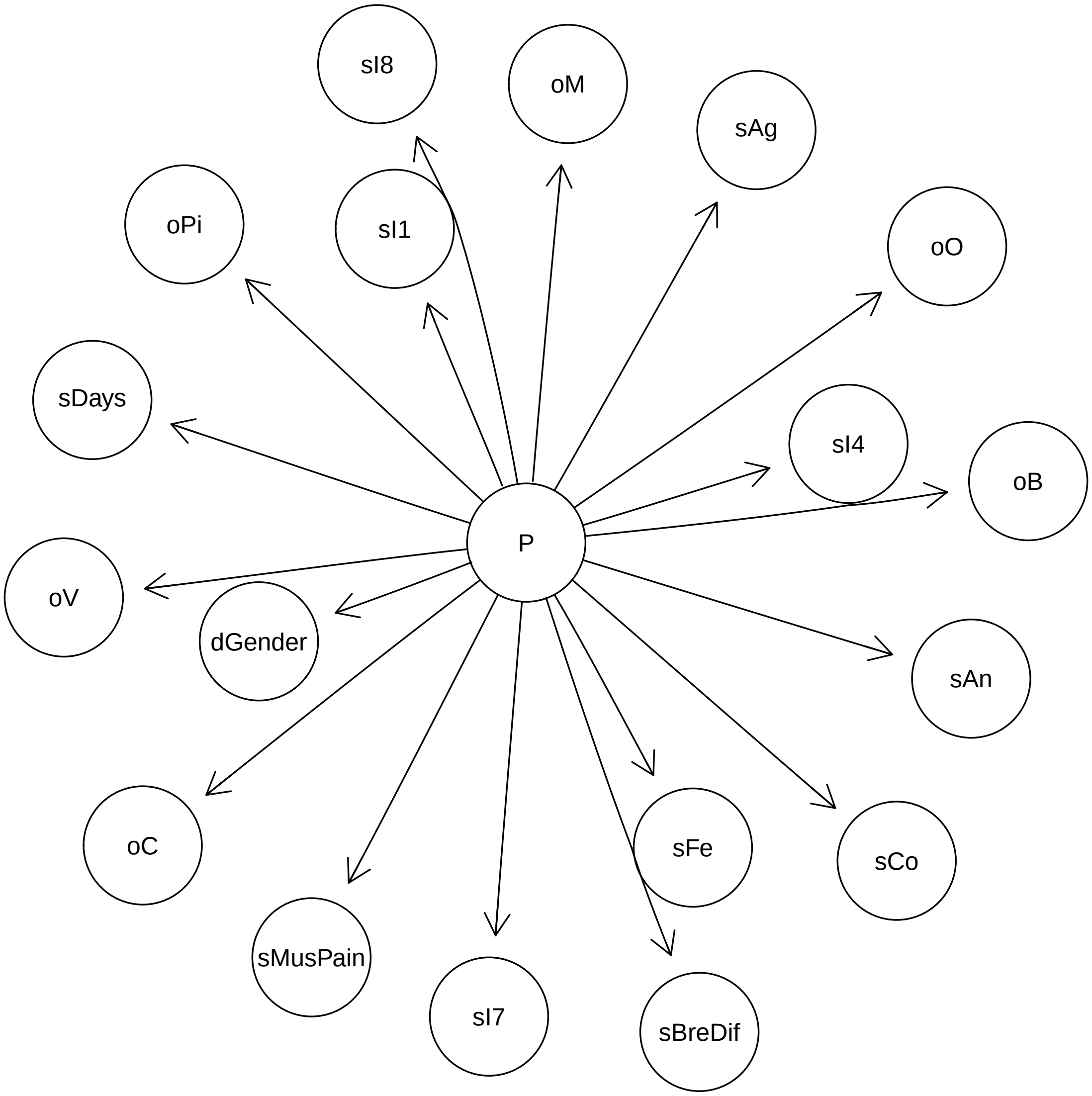


**Figure 1: Structure 1 - Model without Cold.**

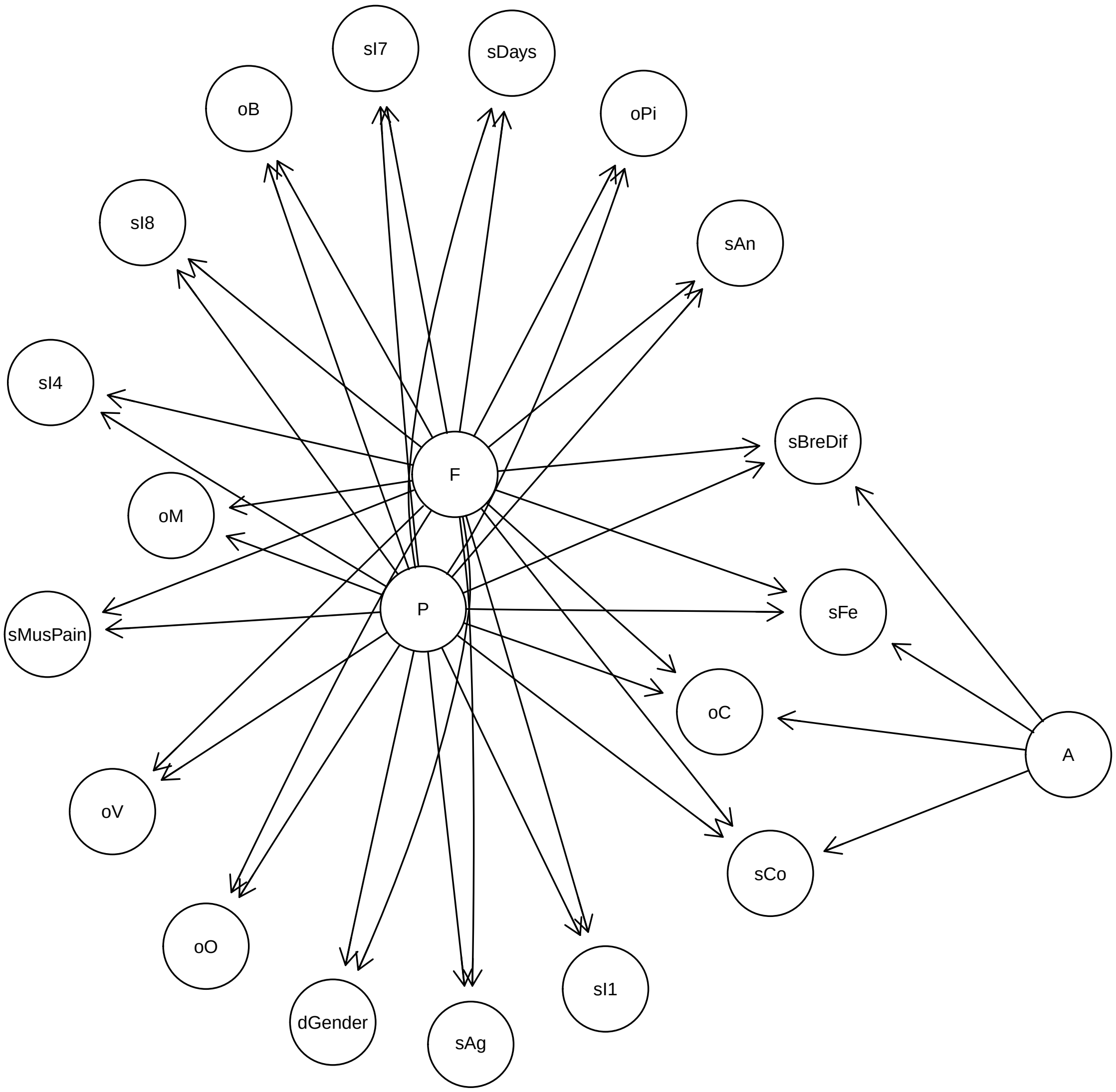


**Figure 2: Structure 2 - Model with Cold and allergies.**

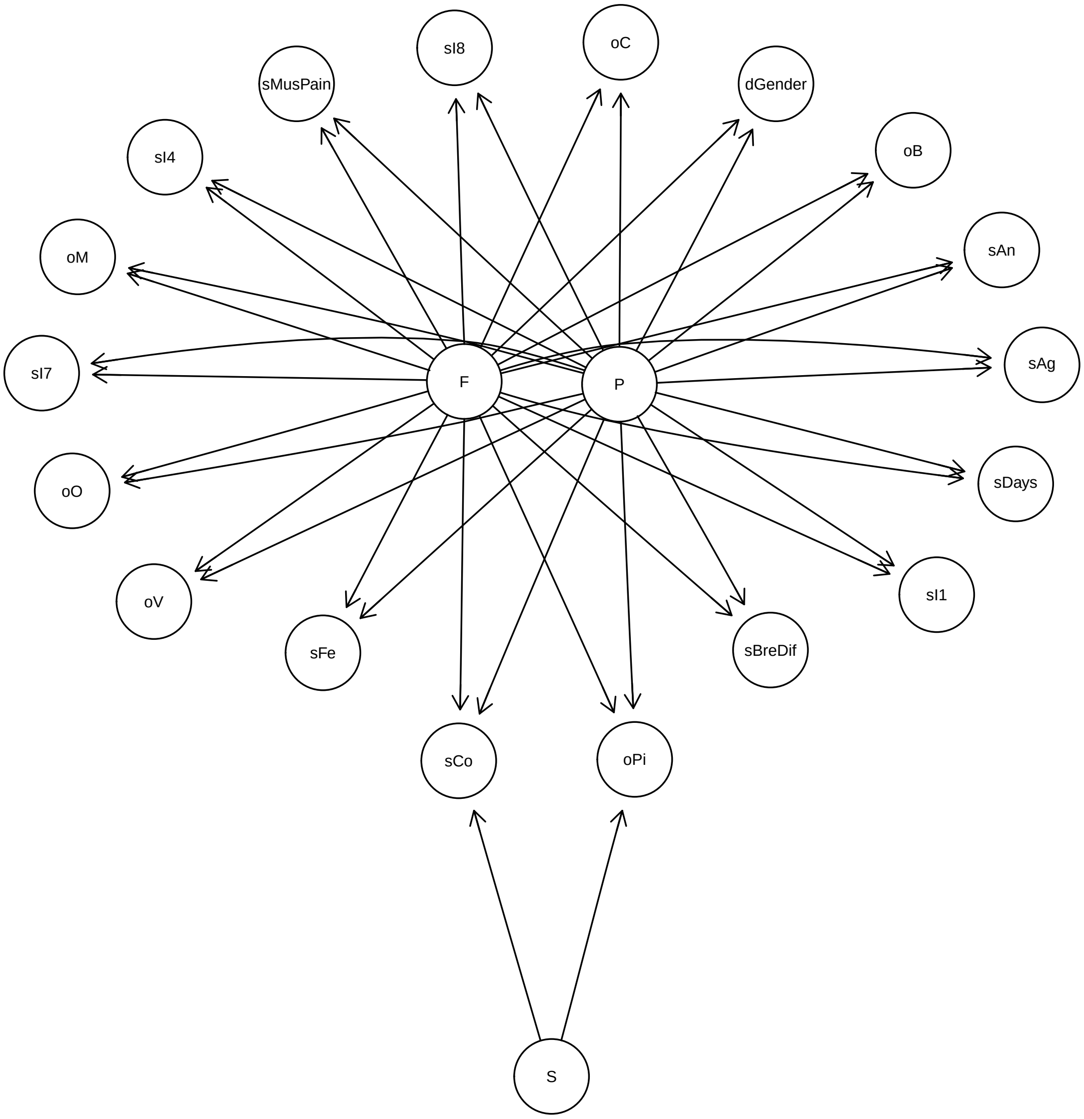


**Figure 3: Structure 3 - Model with Cold and smoking.**

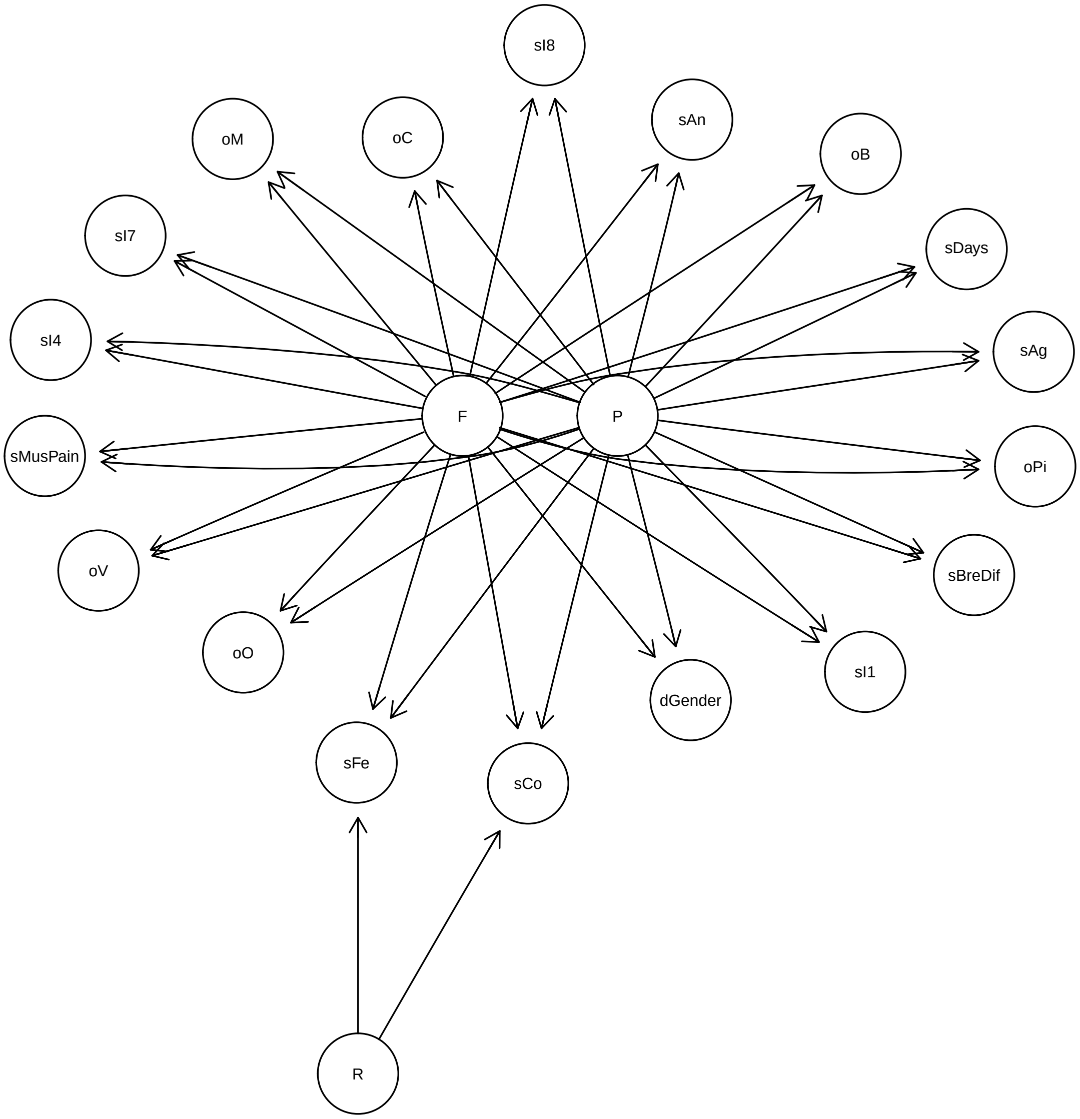


**Figure 4: Structure 4 - Model with Cold and rhinitis.**

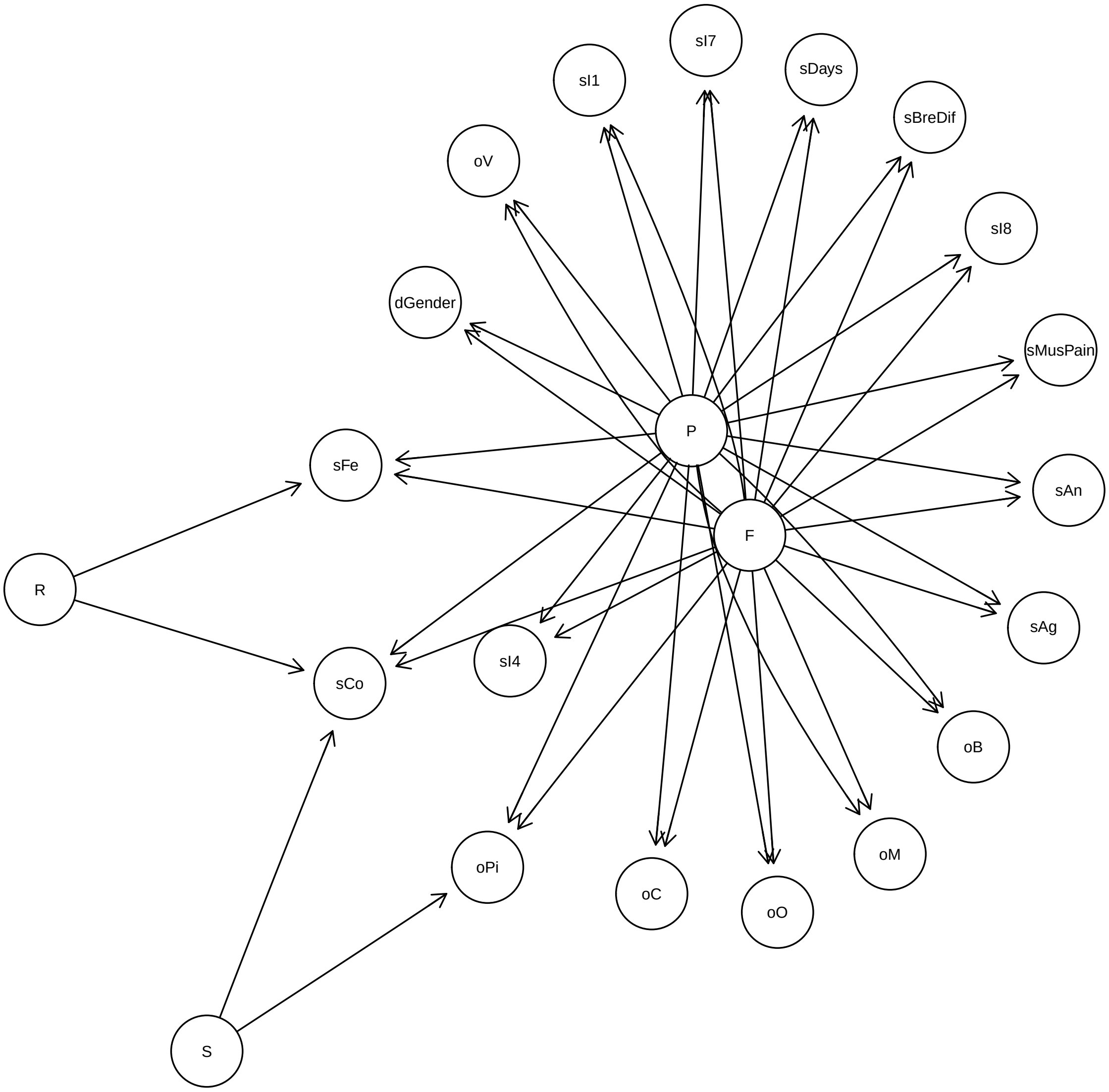


**Figure 5: Structure 5 - Model with Cold, smoking and rhinitis.**

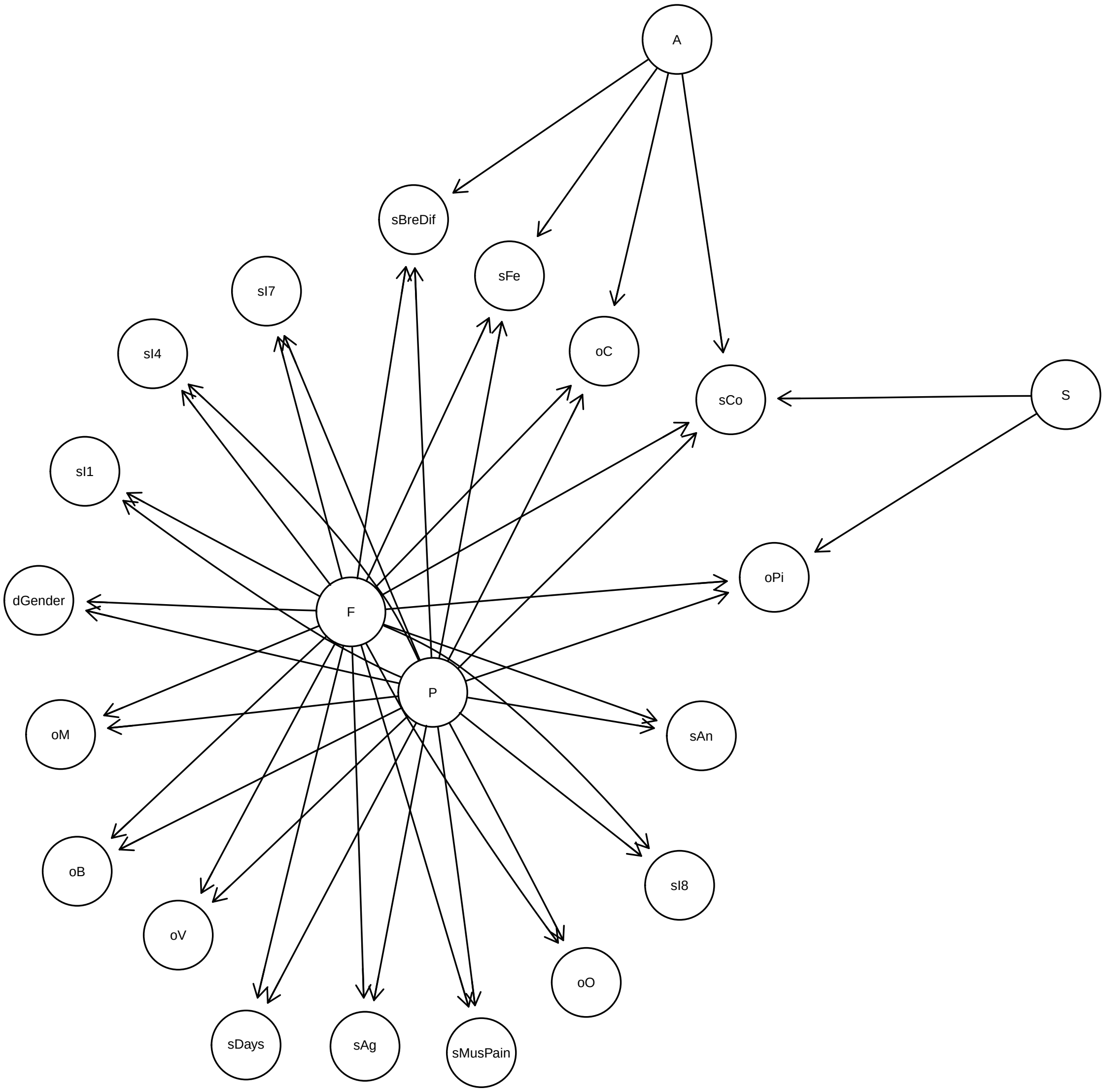


**Figure 6: Structure 6 - Model with Cold, smoking and allergies.**

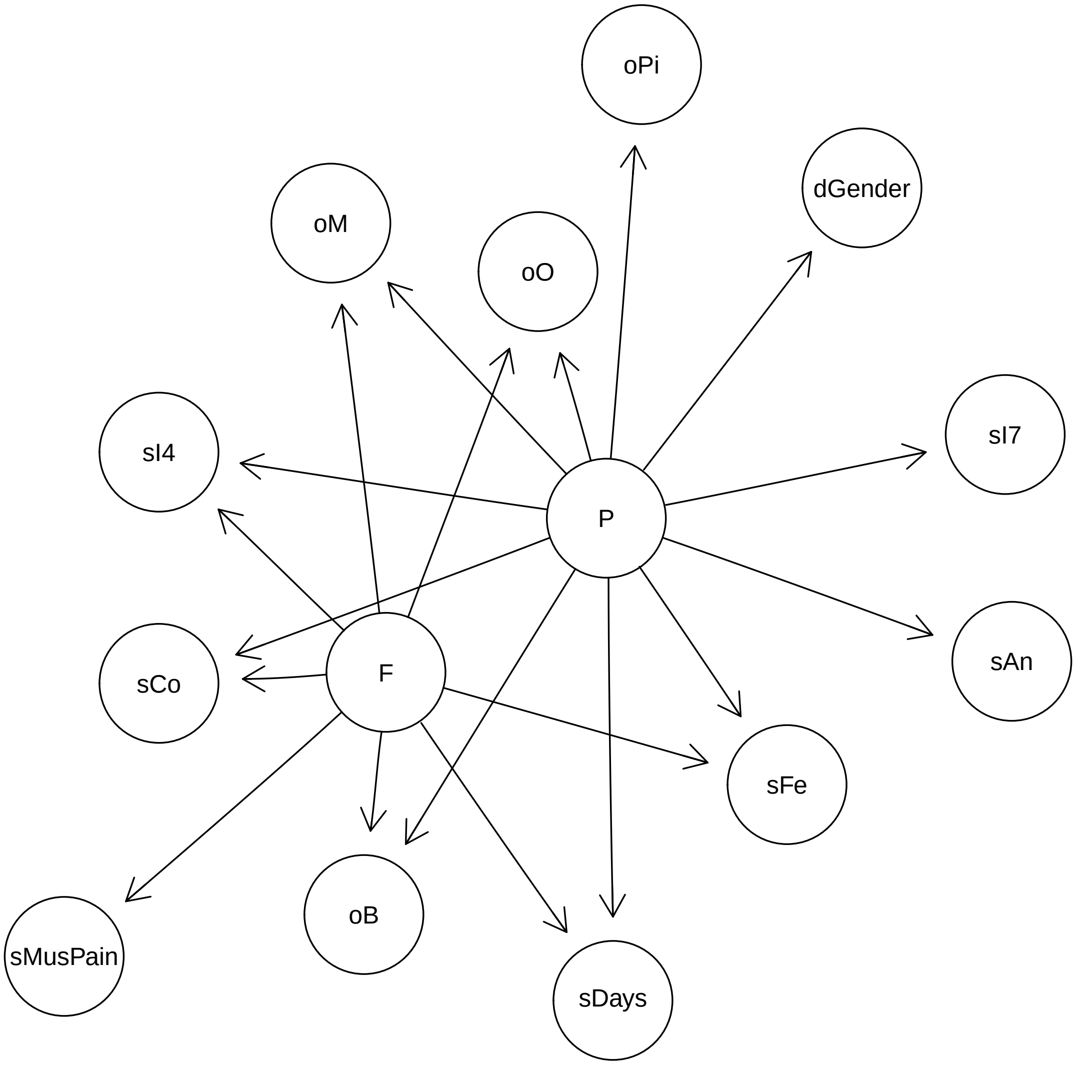


**Figure 7: Structure 7 - Model with Cold and only significant covariates.**

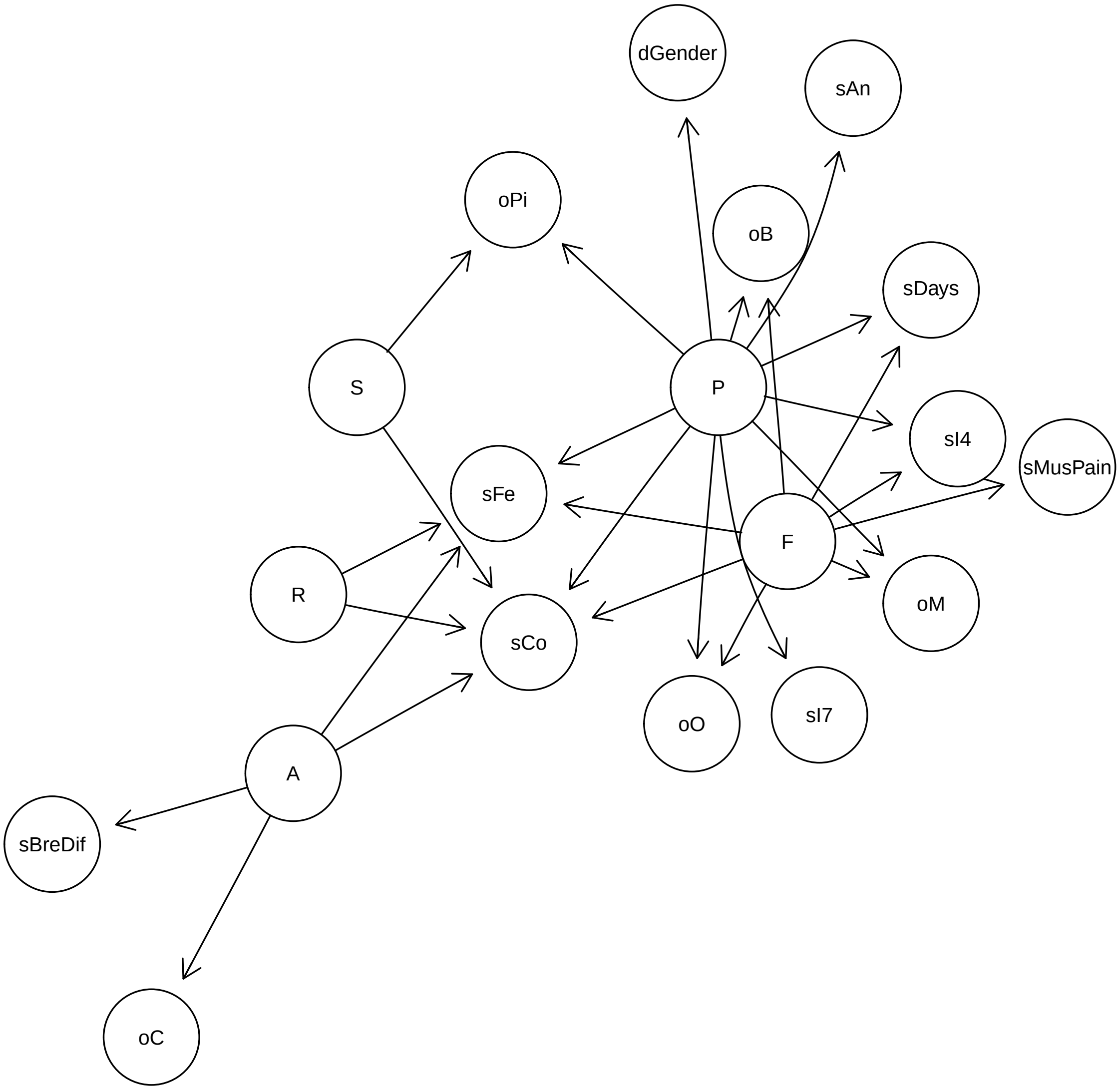


**Figure 8: Structure 8 - Model without Cold and only significant covariates.**

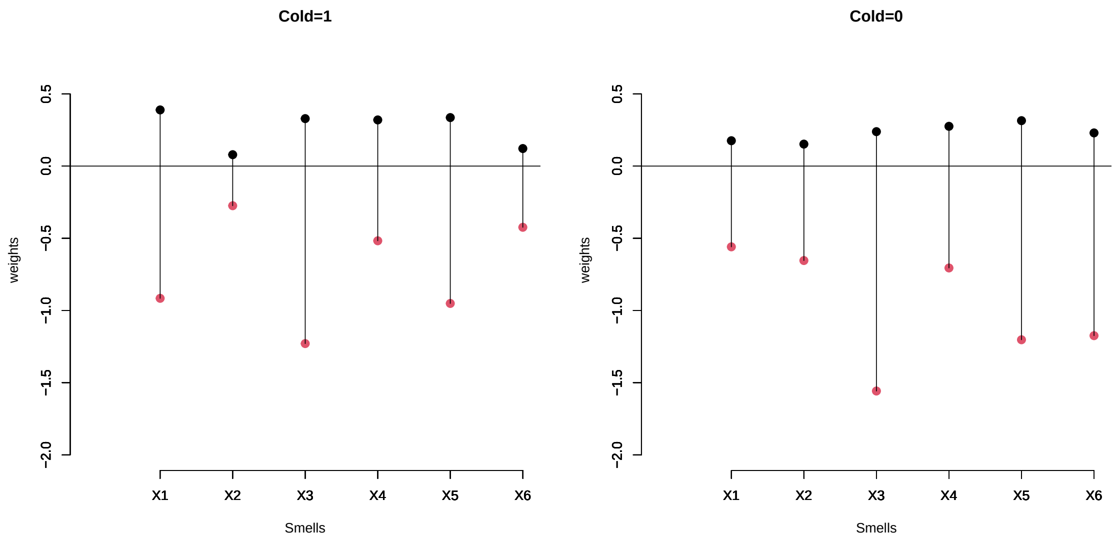


**Figure 9:** Weights contributions of the recognized odors to the risk of a positive PCR result for COVID-19. Red dots represent the weights when the individual fail to recognize an odor, whereas black dots represent the opposite. $X1$ = Banana, $X2$ = Caramel, $X3$ = Mint, $X4$ = Orange, $X5$ = Pineapple, $X6$ = Vanilla

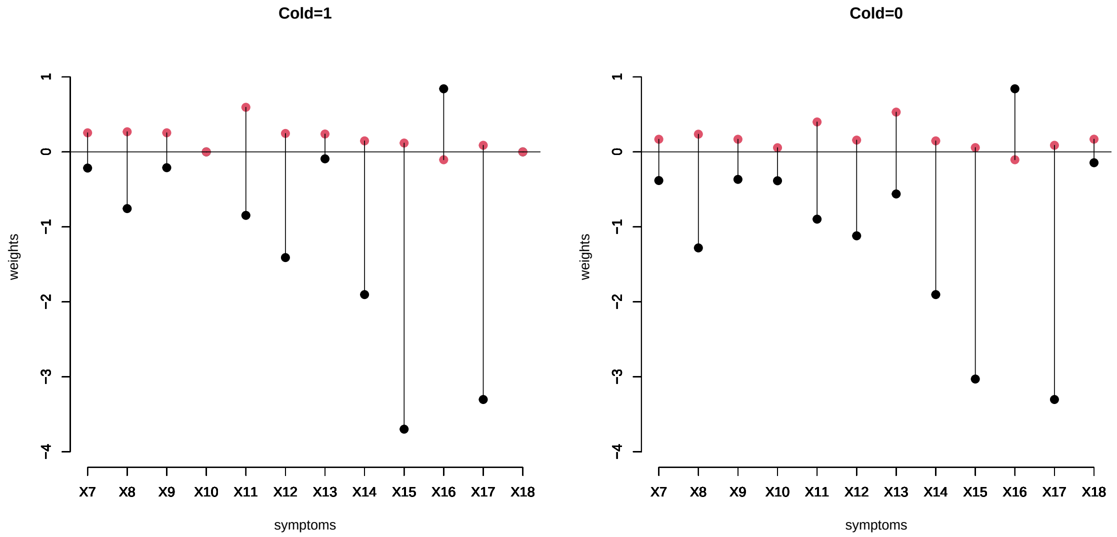


**Figure 10:** Weights contribution of the symptoms to the risk of a positive PCR result for COVID-19. Red dots represent the weights when there is absence of the symptom or condition, whereas black dots represent the opposite, i.e., presence of the symptom or condition. Cough ($X_7$), fever ($X_8$), muscular pain ($X_9$), breathing difficulty ($X_{10}$), self-reported anosmia ($X_{14}$), ageusia ($X_{15}$), five indicator variables ($X_{11}, X_{12}, X_{13}, X_{16}, X_{17}$) and gender $X_{18}$

.

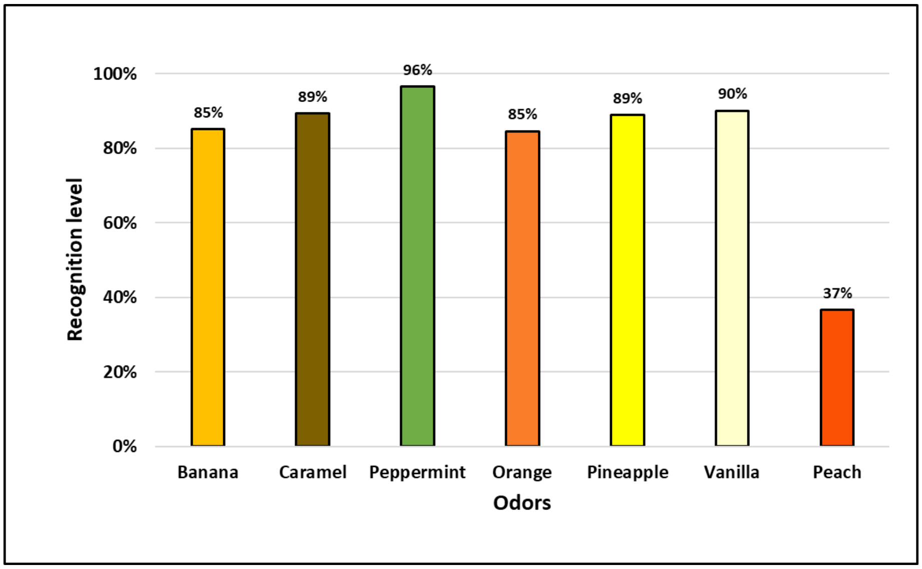


**Figure 11:** Recognition level of 7 odors in a pilot study for the KOR test. All odors were highly recognized in sample population with the exception of peach. The latter odor was excluded from the odor panel of the KOR test.

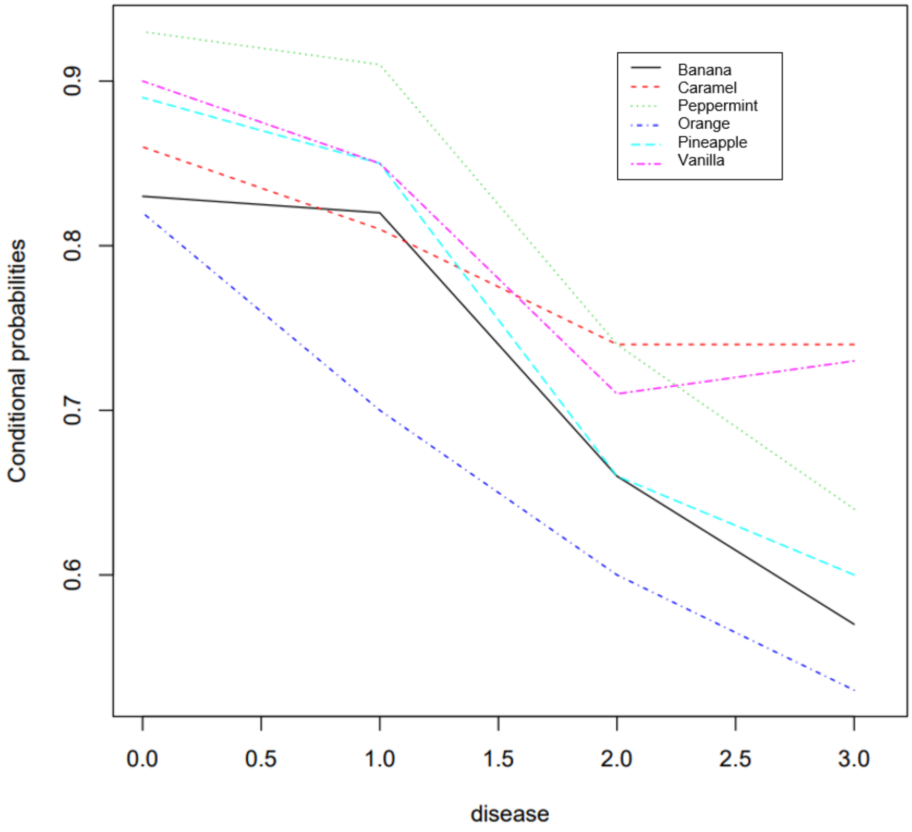


**Figure 12:** Conditional probabilities for the perception of smells given the flu and RT-PCR status: 0: without flu and without COVID-19; 1:with flu and without COVID-19; 2:without flu and with COVID-19; 3:with flu and with COVID-19.

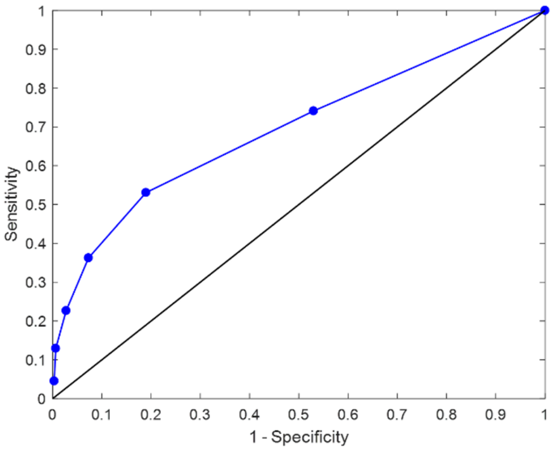


*KOR test intrinsic characteristics for a score of 4.*

| Cutoff k ≤ 4 | | |
|---|---|---|
| Statistic | Value | 95% CI |
| Sensitivity | 53% | 48% to 59% |
| Specificity | 81% | 78% to 84% |
| Positive Likelihood Ratio | 2.8 | 2.3 to 3.4 |
| Negative Likelihood Ratio | 58% | 0.5 to 0.7 |
| Disease Prevalence | 33% | |
| Positive Predictive Value | 58% | 54% to 63% |
| Negative Predictive Value | 78% | 75% to 80% |
| Accuaracy | 72% | |

**Figure 13:** COVID-19 prediction of the olfactory test assuming equal weights for the odors. The best classification performance is obtained with a score of four or less correctly identified odors. The UC-Christus dataset was employed to evaluate the performance of this model.
